# Modelling SARS-CoV-2 transmission in a UK university setting

**DOI:** 10.1101/2020.10.15.20208454

**Authors:** Edward M. Hill, Benjamin D. Atkins, Matt J. Keeling, Michael J. Tildesley, Louise Dyson

## Abstract

Around 40% of school leavers in the UK attend university and individual universities generally host thousands of students each academic year. Bringing together these student communities during the COVID-19 pandemic may require strong interventions to control transmission. Prior modelling analyses of SARS-CoV-2 transmission within universities using compartmental modelling approaches suggest that outbreaks are almost inevitable.

We constructed a network-based model to capture the interactions of a student population in different settings (housing, social and study). For a single academic term of a representative campus-based university, we ran a susceptible-latent-infectious-recovered type epidemic process, parameterised according to available estimates for SARS-CoV-2. We investigated the impact of: adherence to (or effectiveness of) isolation and test and trace measures; room isolation of symptomatic students; and supplementary mass testing.

With all adhering to test, trace and isolation measures, we found that 22% (7% - 41%) of the student population could be infected during the autumn term, compared to 69% (56% - 76%) when assuming zero adherence to such measures. Irrespective of the adherence to isolation measures, on average a higher proportion of students resident on-campus became infected compared to students resident off-campus. Room isolation generated minimal benefits. Regular mass testing, together with high adherence to isolation and test and trace measures, could substantially reduce the proportion infected during the term compared to having no testing.

Our findings suggest SARS-CoV-2 may readily transmit in a university setting if there is limited adherence to nonpharmaceutical interventions and/or there are delays in receiving test results. Following isolation guidance and effective contact tracing curbed transmission and reduced the expected time an adhering student would spend in isolation.

## Introduction

Globally, many countries have employed social distancing measures and nonpharmaceutical interventions (NPIs) to curb the spread of SARS-CoV-2 [1]. In the United Kingdom (UK), the enaction of lockdown on 23rd March 2020 saw the closure of workplaces, pubs and restaurants, and the restriction of a range of leisure activities. The education sector was also impacted, with schools closed (with the exception of children of key workers) and higher education establishments, such as universities, delivering the end of the 2019/2020 academic year via online means.

In the summer months, the national implementation of strict measures transitioned to a localised approach, targeting regions experiencing the highest level of transmission. As the number of SARS-CoV-2 confirmed hospitalised cases and deaths began to decline, many sectors of society cautiously reopened, with measures in place to reduce transmission. Universities began to develop plans to reopen, with several adopting a blended learning strategy of limited face-to-face teaching combined with online lectures. Higher education in the UK comprises a sizeable population of students, with over 2.3 million higher education students enrolled in the 2018/2019 academic year across over 160 higher education providers [2]. This results in a sizeable movement of students nationwide at the beginning and end of academic terms (in addition to international student travel). The migration of students contributes to increased population mobility, which had already grown since the easing of lockdown measures [3, 4]. Thus, there is an associated need for careful management in order to minimise the risk of seeding outbreaks in low prevalence locations.

As of 1st March 2021, the UK had reported in excess of 4 million cases and more than 120,000 COVID-19 deaths [5]. There is, however, a lower risk of severe outcomes in typical student age groups compared with older sections of the population; a higher proportion of cases are expected to be asymptomatic [6], while hospitalisation and mortality rates are lower [7]. In particular, of 83, 529 COVID-19 associated deaths in hospitals reported in England by 1st March 2021, 573 (0.7%) were 20-39 years of age [8].

Nevertheless, the typical contact patterns of students could result in a significant potential to transmit the virus within their social group, amplifying the risk of infection to staff members and those in the local community who may be more vulnerable. Contact studies indicate that students, and in general those aged 20 to 30, report higher numbers of social contacts in their everyday lives compared with other age-groups and occupations [9–11]. In addition, as a consequence of younger age groups more often experiencing asymptomatic infection, there is the prospect of asymptomatically infected students returning home at the end of term and unwittingly transmitting to more vulnerable family members.

The data and science surrounding the SARS-Cov-2 infection is fast moving, so much so that publications can rarely keep pace. This paper was originally written in October 2020, and is a record of the state of our modelling as of that time. At the original time of writing, a small number of modelling analyses had already been carried out pertaining to transmission of SARS-CoV-2 within universities, and subsequent levels of COVID-19 disease [12]. These modelling studies had been predominately US-focused [13–17], potentially due to their earlier return. Paltiel *et al*. [13] modelled the effect of a variety of testing strategies on the number of infections that would arise among 5,000 students during an 80-day semester. Cashore *et al*. [14] and Lopman *et al*. [16] investigated the impact of testing, screening and isolation for Cornell’s Ithaca campus and Emory University in Atlanta, Georgia, respectively.

The size and set-up of UK universities can differ markedly to US counterparts, influencing contact patterns and thus the spread of infection. Though the majority of prior work has not had access to realistic contact structures within the university setting, Brooks-Pollock *et al*. [18] developed a stochastic transmission model based on realistic mixing patterns between students and applied to the University of Bristol. Other UK-centric work has included investigations into the expected number of cases that may be present at the outset of the autumn term in 2020 [19], and a working paper looking at how mathematical approaches may help inform the reopening of higher education spaces to students whilst minimising risk [20].

Many of the previous studies of SARS-CoV-2 outbreaks in a university setting have adopted compartmental modelling approaches, in which individual behaviour and interventions such as contact tracing cannot be readily captured. In this paper, we present an individual-level network-based model framework for transmission of SARS-CoV-2 amongst a student university population, including test, trace and isolation interventions. Contacts occur across household, study and social settings, underpinned by empirical data where possible.

We find that maintaining strong adherence to isolation guidance and engagement in test and trace could both curb the amount of infection throughout the academic term and limit SARS-CoV-2 prevalence at the beginning of the winter break. The use of room isolation offers marginal reductions in the total number of infections, as well as the number of tests administered and time spent in isolation. Mass testing can lead to significant reductions in the total number of infections, though this strongly depends on the adherence of students and the testing procedure implemented. Finally, resource expenditure (including time spent in isolation by students) generally follows a non-monotonic relationship with the strength of and adherence to interventions. Small increases in the latter can lead to increased expenditure. These results show the possible impact of SARS-CoV-2 transmission intervention measures that may be enacted within a university population during the forthcoming academic year.

## Methods

We adopted a network approach to capture the interactions between students in different settings, upon which we ran an epidemic process. In this section we provide details regarding the (i) network generation, (ii) data sources used to parameterise the network contact structure, (iii) model for SARS-CoV-2 transmission and COVID-19 disease progression, and (iv) simulation protocol used to assess the scenarios of interest.

### Network model description

Our network model comprised four layers: (i) households, (ii) study groups/cohorts, (iii) organised societies and sports clubs, and (iv) dynamic social contacts.

#### Household contact layer

We considered contact networks on-campus and off-campus separately. The network for on-campus accommodation contained a hierarchical structure, from the smallest scale to the largest: household (typically based around a shared kitchen), floor, block and hall. We constructed the on-campus accommodation units to match that of a representative campus based university. We assigned students resident off-campus to households with sizes sampled from an estimated student household size distribution (see Supporting Text S1).

Within a household, irrespective of on-campus or off-campus location, we assumed a fully connected network. Thus, every member of the household is assumed to be in regular contact with every other member, with no variability between members. In addition, on-campus students could make contact with other students situated in the same floor or accommodation block. These contacts were randomly generated each day with a fixed probability (this probability is greater for students within the same floor, compared to the same block; see Table 1).

**Table 1:** Description of network contact parameters.

| Description | Degree distribution | Source |
| --- | --- | --- |
| Household (static) | Fully connected | Assumption |
| Within accommodation block (on-campus only) | Floor contact: daily contact probability of 0.1. Block contact: daily contact probability of 0.05. | Assumption |
| Study cohort (undergraduate) | Log-normal(1.646,1.590) | Fitted from Social Contact Survey [9–11] |
| Study cohort (postgraduate) | Log-normal(1.211,1.128) | Fitted from Social Contact Survey [9–11] |
| Societies & sports clubs | Probability of link with each group member: 0.05 for societies, 0.1 for sports clubs | Assumption |
| Dynamic social (undergraduate) | Log-normal(1.748,1.331) | Fitted from Social Contact Survey [9–11] |
| Dynamic social (postgraduate) | Log-normal(1.223,1.125) | Fitted from Social Contact Survey [9–11] |

#### Study group/cohort contact layer

We assumed that the enforcement of COVID-secure measures within the classroom was sufficient to prevent any transmission between students in that setting. However, the study group/cohort contact layer does account for the fact that students in the same classes are likely to regularly study and socialise together outside of the classroom. To construct this layer, we partitioned the student population into 84 cohorts based on department and stage of study: first year undergraduate, non-first year undergraduate, and postgraduate (see Supporting Text S2). To generate the contacts made within each cohort, we used a configuration model [21] to allow the specification of a desired degree distribution.

#### Contacts in organised societies and sports clubs

A prominent aspect of the university experience is the presence of societies and sports clubs. Each student was allocated to between 0 and 5 such groups (further details in the next section). To construct contacts within these groups, we applied a constant probability of forming a contact with each other individual in the group, with that probability differing based on whether the group was a society or sports club. These links did not alter during the course of a simulation. We assumed each social group met on three fixed days each week of the academic term (meeting schedules could differ between groups), with all members attending all meetings.

#### Dynamic social contacts

In this layer, we captured random, dynamic contacts made each day with any other individual in the student population. Each day, random connections were selected for each student according to a specified distribution, dependent on their level of study (Table 1).

### Contact parameterisation

We characterised the network structure across the various contact layers by applying two different approaches.

The first method was a data-driven approach, using data from the Social Contact Survey [9–11]. The Social Contact Survey was a paper-based and online survey of 5,388 participants in the United Kingdom, conducted in 2010. We extracted records provided by 341 students, with a total of 10,275 contacts. These data informed the network construction parameters for the study group/cohort and dynamic social contact layers. We divided the data according to level of study: undergraduate (282 students) or postgraduate (59 masters and PhD students). Part-time, remote and mature students were excluded. We fit parameters for these contact distributions using maximum likelihood estimation via the fitdistrplus package in R.

The second method was a subjective approach, used when we did not have relevant data available to parameterise the given contact layer. This was applied to the formulation of random contacts within on-campus accommodation blocks and organised social club contacts. We provide a summary of the network parameterisation in Table 1.

#### Study group/cohort contacts

We used student contacts recorded in the Social Contact Survey, restricted to those listed as occurring in a work or school setting. We kept entries specifying a duration of 60 minutes or more, occurring more than once per week. We assumed that the retained contacts with these characteristics would be reflective of contacts made in a classroom / study group context. There were a total of 135 students with 2497 relevant contacts (111 undergraduates contributed 2309 contacts, 24 postgraduates contributed 188 contacts). We fitted log-normal distributions for undergraduates and postgraduates independently, using a mean and standard deviation parameterisation, acquiring distributions of Log-normal(1.646, 1.590) and Log-normal(1.211, 1.128), respectively (Fig. 1(a)).

**Fig. 1:**
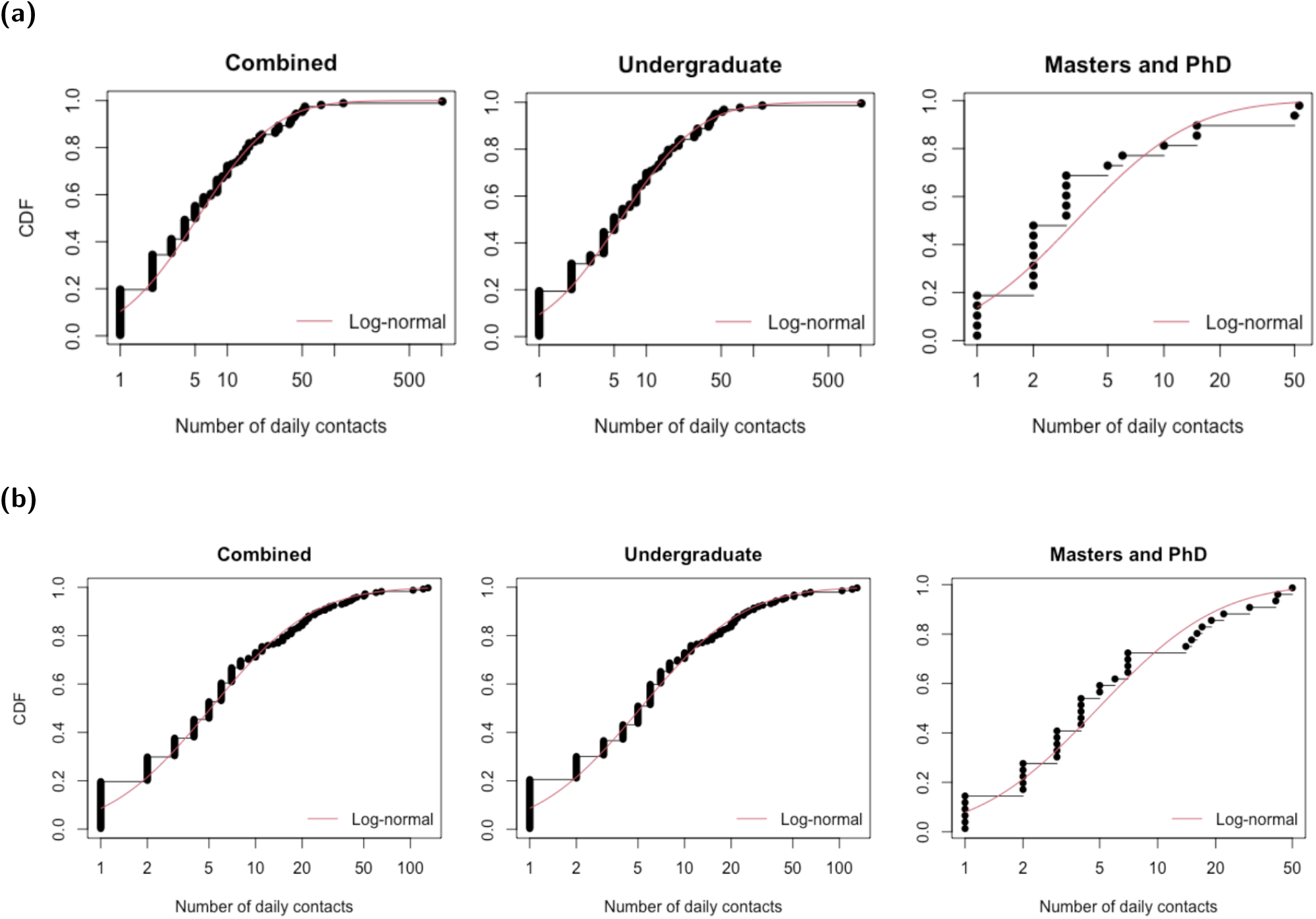
Cumulative distribution functions for the number of daily contacts for all students, undergraduates and postgraduates. Black dots and lines depict the empirical data. The red solid line corresponds to the best-fit log-normal distribution. **(a)** Daily cohort contacts using a total of 135 students with 2497 relevant contacts (111 undergraduates contributed 2309 contacts, 24 postgraduates contributed 188 contacts). **(b)** Daily social contacts, using a total of 206 students with 2249 relevant contacts (168 undergraduates contributed 1888 contacts, 38 postgraduates contributed 361 contacts).

#### Dynamic social contacts

We used student contacts recorded in the Social Contact Survey, excluding those occurring at home and those occurring for the first time. Furthermore, we limited valid contacts to those recorded as either involving touch or lasting longer than 10 minutes. Finally, valid contacts had to last less than 60 minutes, as longer duration contacts would be captured by the cohort and society contact layers. A total of 206 students with 2249 relevant contacts were included (168 undergraduates contributed 1888 contacts, 38 postgraduates contributed 361 contacts).

Overall, undergraduates and postgraduates had a very similar number of social contacts (daily medians of 5.19 and 4.9 respectively). We fitted a Log-normal(1.748, 1.331) distribution for undergraduates, and Log-normal(1.223, 1.125) for postgraduates (Fig. 1(b)).

#### Random on-campus accommodation contacts

Given the limited data available to parameterise a degree distribution for these contacts, we took a pragmatic approach and assumed a low constant probability of contacts occurring in the broader accommodation unit. We assumed these contact probabilities lessened for higher levels of accommodation hierarchy. Specifically, we attributed a higher chance of interacting with someone on the same floor (daily chance of contact of 10% per student with each other student) than someone on another floor within the same block (daily chance of contact of 5%). We assumed no random contacts at the highest accommodation levels, i.e. between separate blocks of the same hall, or between different halls.

#### Contacts in organised societies and sports clubs

We did not have the necessary information to parameterise contacts within this layer using a data-driven approach. Therefore, we stress that the values stated here are subjective and alternative proposals would add to result variability.

We considered a community of 335 organised social groups, comprising 270 societies and 65 sports clubs. We allowed a breadth of group sizes, randomly assigning each group a membership size of 10 to 100 (in increments of 10). We chose a monotonically decreasing probability mass function for the number of groups each student actively participated in: 50% of students not in any group; 40% involved in a single group; 2.5% for each of two, three, four and five groups.

Following group assignment, we established contacts between members with a fixed probability of each link existing. We set these probabilities to 0.05 for societies and 0.1 for sports clubs. Accordingly, a student was likely to make more contacts in groups with large membership. For simplicity, we retained the same arrangement of contacts for each meeting.

### Epidemiological model

#### Disease states

We ran a susceptible-latent-infectious-recovered (*SEIR*) type disease process on the network structure. Once infected, we assumed infectiousness could start from the following day. We assumed an Erlang-distributed incubation period, with shape parameter 6 and scale parameter 0.88 [22].

The distribution of infectiousness had a four day pre-symptomatic phase, followed by a ten day symptomatic phase. This gave a total of 14 days of infectivity and a minimum 15 day infection duration. The infectiousness temporal profile weighted the contact setting transmission risk (see the subsequent subsection on *Setting transmission risk*) across the duration of the infectious period (for the full temporal profile, see Table 2). It was based on a Gamma(97.2, 0.2689) distribution, with shape and scale parameterisation, shifted by 25.6 days [23, 24]. Following completion of the infectious period, the individual entered the recovered state (see Fig. S7 for a schematic representation of the model).

**Table 2:** Description of epidemiological parameters. Note the transmission risk parameters correspond to the probability of transmission (to a susceptible individual) over the entire duration of the infectious period for a symptomatic case.

| Description | Distribution | Mean | Source |
| --- | --- | --- | --- |
| Incubation period | Erlang(6,0.88) | 6.82 days | [22] |
| Infectiousness profile | Infectivity profile over 14 days:<br>[0.0369, 0.0491, 0.0835, 0.1190,<br>0.1439, 0.1497, 0.1354, 0.1076,<br>0.0757, 0.0476, 0.0269, 0.0138,<br>0.0064, 0.0044] | N/A | [23, 24] |
| Proportion of cases asymptomatic | Uniform(0.5,0.8) | 0.65 | [25, 26] |
| Relative infectiousness of an asymptomatic | Uniform(0.3,0.7) | 0.5 | [27, 28] |
| Household contact transmission risk (household size 2, 3, 4, 5+) | Size 2: Normal(0.48,0.06)<br>Size 3: Normal(0.40,0.06)<br>Size 4: Normal(0.33,0.05)<br>Size 5+: Normal(0.22,0.05) | 0.48<br>0.40<br>0.33<br>0.22 | Assumption |
| Society contact transmission risk | Normal(0.12,0.018) | 0.12 | Assumption |
| Class contact transmission risk | Normal(0.0,0.0) | 0.00 | Assumption |
| All other non-household contact transmission risk | Normal(0.25,0.035) | 0.24 | Assumption |

#### Asymptomatic transmission

Infected individuals could be either asymptomatic or symptomatic, according to a specified asymptomatic probability. There remains significant uncertainty as to what this probability should be, however community surveillance studies informed this parameter. The REal-time Assessment of Community Transmission-1 (REACT-1) study found approximately 70% of swab-positive adults and 80% of swab-positive children were asymptomatic at the time of swab and in the week prior [25]. Note that this includes presymptomatic infected individuals who would later go on to display symptoms. This fell to 50% at later stages of the study [26]. To reflect this uncertainty, for each simulation we sampled the asymptomatic probability from a Uniform(0.5, 0.8) distribution.

There remains limited data available to provide a robust quantitative estimate of the relative infectiousness of asymptomatic and symptomatic individuals. However, there are indications that asymptomatic individuals could be less infectious than symptomatic individuals [27, 28]. Therefore, we assumed that asymptomatic individuals had a lower risk of transmitting infection compared to symptomatic individuals. To reflect the uncertainty in this area, for each simulation we sampled the relative infectiousness of asymptomatics compared to symptomatics from a Uniform(0.3, 0.7) distribution. This was sampled independently to the asymptomatic probability. The sampled value was applied as a scaling on transmission risk, applied evenly throughout the duration of infectiousness (*i*.*e*. with no time dependence, see the subsection *Probability of transmission per contact*).

#### Setting transmission risk

Attributing risk of transmission to any particular contact in a particular setting is complex, due to the huge heterogeneity in contact types. We used a data-driven approach to obtain the relative risk of transmission within each network layer. We then scaled these risks equally in order to obtain an appropriate growth rate of the disease.

For each contact setting (network layer), the transmission risk corresponded to the probability of a susceptible individual being infected over the course of the entire infectious period for an infected with a relative infectiousness of 1, assuming the susceptible and infectious individual were in contact in the specified setting every day. The transmission risk was then scaled to obtain the probability of transmission occurring across a susceptible-infectious contact pair on a given day (see the subsection *Probability of transmission per contact*).

For household transmission, we used estimates of adjusted household secondary attack rates from a UK based surveillance study [29]. We attributed a household secondary attack rate to each student based on the size of their household. We sampled the attack rates from a normal distribution with mean dependent on the household size: 0.48 for a household size of two, 0.40 for three, 0.33 for four, and 0.22 for five or more. The standard deviation of the normal distribution for households of size two or three was 0.06, and for households of four or more was 0.05.

For transmission risk in other settings, we performed a mapping from the Social Contact Survey [9–11] to obtain a relative transmission risk compared to household transmission. To obtain the means, we used the central estimate of adjusted household secondary attack rate for those aged 18-34 of 0.34 [29] and scaled this based on the characteristics of contacts in different locations, obtained from the contact survey (further details in Supporting Text S3). Standard deviations were set to have a constant size relative to the mean. Transmission risks were consistent across all non-household settings, except within organised societies where we assigned a lower transmission risk to reflect the implementation of COVID-secure measures that would be required to permit these meetings to take place. We also reiterate that we attributed zero transmission risk to face-to-face classes, for the same reason.

To calibrate the relative transmission risks to achieve an uncontrolled reproductive number, *R*_*t*_, in the expected range of 3 − 4, we applied an equal scaling of 0.8 to all of the transmission risks calculated above (see Supporting Text S4).

#### Probability of transmission per contact

We outline here how the setting transmission risk, infectiousness temporal profile and relative infectiousness of an individual were used to compute the probability of transmission across an infectious-susceptible contact pair.

For an infectious individual *j* on day *t* of their infectious state, the probability of transmission per susceptible contact in contact setting *s*, denoted *p*_*j,s*_(*t*), was given by the product of four components:

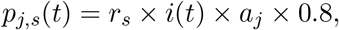

with *r*_*s*_ the transmission risk in setting *s, i*(*t*) the value of the infectiousness temporal profile on day *t* (Table 2), *a*_*j*_ the relative infectiousness of individual *j* (taking either value 1 for cases that were symptomatic during the infection episode, or the sampled value for relative infectiousness of asymptomatics compared to symptomatics otherwise), and 0.8 the scaling applied to calibrate the system to achieve (in the majority of simulations) an *R*_*t*_ in the range of 3-4 for the initial phase of the outbreak.

### Isolation, test and trace

#### Testing and isolation measures

Upon symptom onset, an adhering infected student would immediately take a test and enter isolation for 10 days. At that time, their household would also enter self-isolation for 14 days (matching the UK government guidance prior to 14th December 2020, when self-isolation for contacts of people with confirmed coronavirus was shortened from 14 days to 10 days across the UK) [30]. Isolation was assumed to remove all non-household contacts for the period of isolation.

We assumed that an isolating student would remain in isolation for the required amount of time, or until a negative test result was returned. We included a two day delay between taking the test and receiving the result. We assumed the test had 100% specificity and its sensitivity was dependent upon time since infection (we used the posterior median profile of the probability of detecting infection reported by Hellewell *et al*. [31]).

In the event that the test result from the index case was negative, household members would be released from isolation, as long as no other members had become symptomatic during that time. The index case remained in self-isolation if they had independently been identified via contact tracing as a contact of a known infected; otherwise, that student also left self-isolation.

#### Forward contact tracing

The modelled tracing scheme looked up contacts for an index case up to five days in the past. It was assumed that tracing took place on the third day after symptom onset, following testing and a two day delay to return a positive result. Thus contacts may be recalled up to two days prior to the onset of symptoms. We assumed that a student would be able to recall all of their regular contacts for that time. However, we assumed that the probability of a student being able to recall their ‘dynamic’ contacts diminished with time, from 0.5 one day previously, reducing in increments of 0.1, such that the probability of successfully tracing a contact five days in the past is 0.1. Once again, other assumptions could be explored and a wider range of assumptions, collectively, would generate more variation in the results.

Contacts of a confirmed case were required to spend up to 14 days in self-isolation [32] (matching the UK government guidance prior to 14th December 2020, when self-isolation for contacts of people with confirmed coronavirus was shortened from 14 days to 10 days across the UK). We set the isolation period to elapse 14 days after the index case became symptomatic.

We give an overview of isolation, test and trace related parameters in Table 3.

**Table 3:**
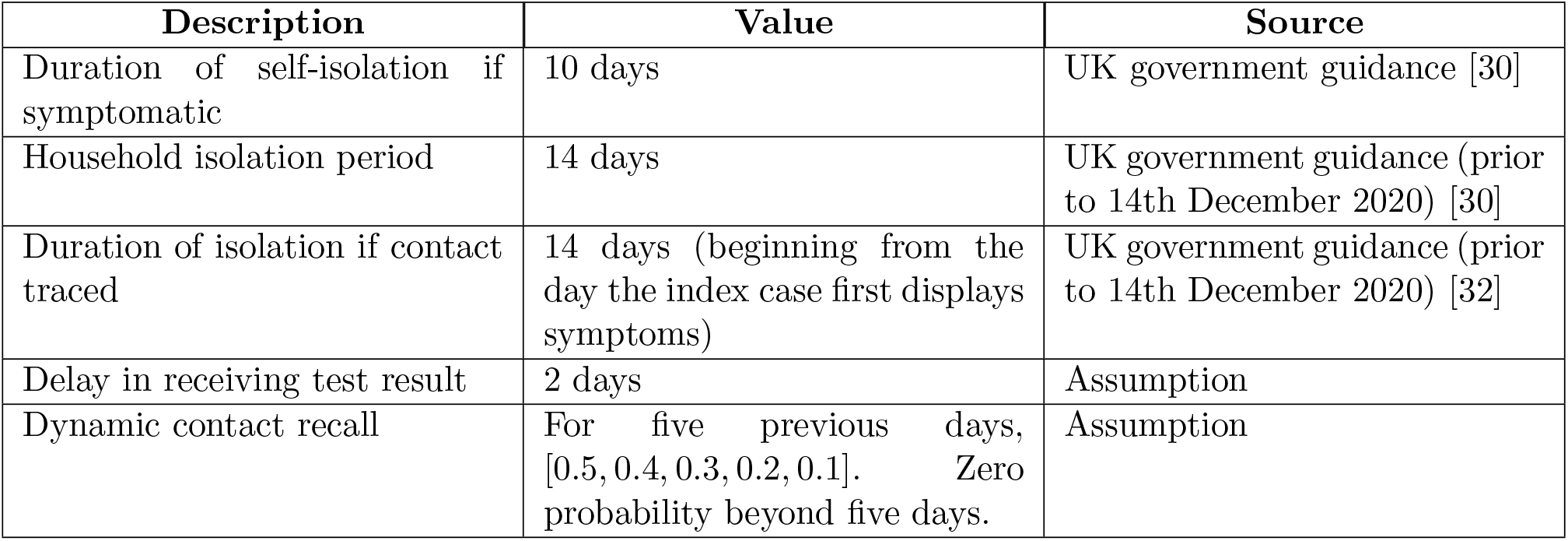
Description of isolation, test and trace related parameters.

### Simulation outline

We used this model framework to evaluate the transmission dynamics of SARS-CoV-2 amongst a university student population during the autumn term of the 2020/2021 academic year, and the potential impact of both adherence to the guidance and additional interventions.

We ran all simulations with an overall student population of 25,000, with 7,155 students resident on-campus and the remainder off-campus. Simulation time corresponded to 77 days, encompassing the length of welcome week plus the ten week academic term.

We seeded the number of latent, asymptomatic and recovered individuals based on UK regional prevalence estimates for 26th September 2020 and student flow data (we provide further methodological details in Supporting Text S5). We assumed there were no symptomatic infected students present at the beginning of each simulation.

Unless stated otherwise, for each parameter configuration we ran 1,000 simulations, amalgamating 50 batches of 20 replicates; each batch of 20 replicates was obtained using a distinct network realisation. We performed the model simulations in Julia v1.4 - 1.5. Code for the study is available at https://github.com/EdMHill/covid19_uni_network_model.

Our analysis comprised of three strands, assessing the impact of: i) adherence to isolation requirements and engagement with test and trace, ii) adopting a policy of strict room isolation for on-campus residents displaying symptoms, and iii) mass testing. In each area of analysis, we reported various measures relating to the prevalence of infection and expenditure of resources due to intervention. The latter includes, and makes a point of, the time spent in isolation by students. We outline each of the three strands in further detail below.

### Adherence to isolation, test and trace

We varied adherence to isolation from no adherence (value 0) to full adherence (value 1) at increments of 0.1. This value was treated as the probability that a student adheres. Whether or not each student will adhere to guidelines was randomly set at the start of each simulation and remained fixed for the duration of that simulation.

Under full adherence, every student who was required to isolate would do so. Isolation was required if a student had symptoms, was in the same household as someone with symptoms, or had been identified as a contact of an infected individual by contact tracing. Additionally, every student would also engage with test and trace. Under no adherence, no students would isolate for any reason, or engage with test and trace. For intermediate values of adherence, an adhering student would adhere to all isolation requirements and engage with testing and tracing, whereas a non-adhering student would adhere to no isolation requirements and would not engage with test and trace. As such, there was no partial adherence to measures by individual students.

### Use of room isolation

For those resident in on-campus accommodation and suffering from COVID-like symptoms, another applicable intervention may be the use of room isolation. Successful implementation of such an intervention would require the student to remain in their room at all times. Sufficient support for the isolating students is vital, including the delivery of meals and other essentials, as well as rehousing those students that reside in accommodation with communal bathrooms (those in rooms with en-suite bathrooms would be able to isolate in their own rooms).

We modelled this intervention by assuming those put into room isolation had no contacts. This extended the standard isolation measures, which still allowed contacts within households. We applied these measures on the same day that the student reported being symptomatic. We assumed students stayed in room isolation until the end of their symptomatic period, irrespective of the test result received.

### Mass testing

We explored the implementation of mass testing, varying the timing and frequency of tests: a single instance on day 21 (end of week 2 of the academic term); a single instance on day 63 (end of week 8 of the academic term); regular mass testing every two weeks (on the first Monday of each fortnight); regular mass testing on a weekly basis (on the Monday of each week). Additionally, we varied coverage amongst the eligible student population: all students, on-campus resident students only, and off-campus resident students only. We carried out sensitivity to the underlying adherence to isolation measures by performing this analysis for adherence probabilities of 0.2 (low), 0.5 (moderate) and 0.8 (high), respectively.

Students who had previously reported infection and subsequently received a positive test were excluded from mass testing. We also assumed that all tests were performed on a single day and those who received a positive test result were immediately placed into room isolation. Contact tracing was performed rapidly such that those contacts who were both traceable and adhered to isolation guidance were isolated from the next day.

In a mass testing campaign, the test’s ability to correctly identify asymptomatic infections is an additional source of uncertainty. While symptomatic and asymptomatic individuals have similar average peak viral loads and proliferation stage duration, the average duration of their clearance stages has been observed to differ [33, 34]. Since the probability of testing positive is likely a function of viral load, this suggests that symptomatic and asymptomatic test sensitivity may differ. We assumed that the probability of asymptomatic individuals testing positive was equal to that of symptomatic individuals until the peak of infection. However, after the peak it was assumed to decay more rapidly, such that the probability of an asymptomatic individual testing positive 6.7 days after the peak would equal the probability of a symptomatic individual testing positive 10.5 days after the peak (corresponding with findings from Kissler *et al*. [33], who estimated an average duration of clearance of 10.5 days in symptomatic cases versus 6.7 days in asymptomatic cases). However, we highlight that this is an area of considerable uncertainty. Future studies detailing the testing probability of asymptomatic individuals, and the specific relationship between viral load and testing probability, would be a valuable contribution to this area.

## Results

### Adherence to isolation, test and trace

We assessed how changes in adherence to isolation, test and trace interventions affected the spread of the virus within the student population over the autumn term. We reported measures pertaining to both the level of infection and the resources expended in implementing the interventions (including time spent by students in isolation).

As adherence to isolation, test and trace increased, we observed a lower number of total infections (the sum of both identified cases and undiagnosed infections; Fig. 2(a), Table S3). Specifically, with no adherence (equivalent to the absence of interventions), we estimated a median proportion of 0.69 (95% prediction interval: 0.56-0.76) of the entire student population would be infected during the term. In contrast, with full adherence, the median proportion infected was 0.22 (95% prediction interval: 0.076-0.41). Irrespective of adherence, the proportion of students infected was consistently higher for those living in on-campus accommodation compared to off-campus.

**Fig. 2:**
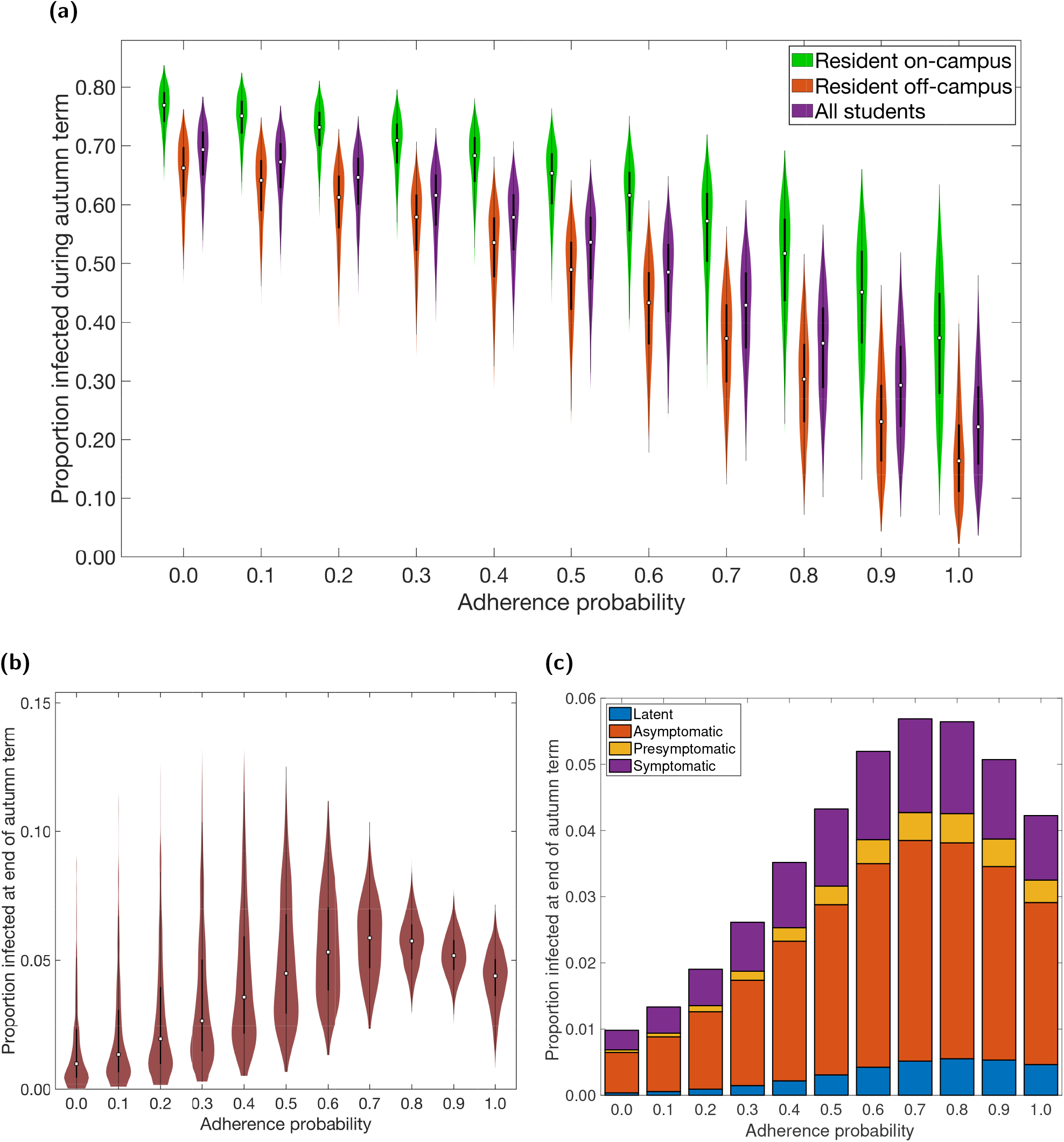
Infection associated epidemiological measures over the autumn term under differing levels of adherence to NPIs. Outputs summarised from 1,000 simulations (with 20 runs per network, for 50 network realisations) for various levels of adherence to NPIs. In all the violin plots, the white markers denote medians and solid black lines span the 25th to 75th percentiles. **(a)** Over the duration of the autumn term, distributions relative to students resident on-campus only (green violin plots), students resident off-campus only (orange violin plots) and to the overall student population (purple violin plots) for proportion infected. Maintenance of nonpharmaceutical interventions and effective contact tracing curbed transmission. On-campus residents were more likely to become infected compared to students living off-campus. For percentile summary statistics, see Table S3. **(b)** Proportion of students infected at the end of the autumn term. **(c)** Under each level of adherence, we display the median proportion of the student population in latent (blue), asymptomatic (orange), presymptomatic (yellow), symptomatic (purple) infected states at the end of the autumn term; for a given adherence value the height of the bar in panel (c) corresponds to the median point in panel (b). For percentile summary statistics, see Table S4.

Although increased adherence resulted in fewer infections overall, this was not necessarily true for the number of infections present at the end of term. We found that for adherences between 0-0.7, increasing adherence resulted in a greater number of infected students at the end of term, rising from a median estimate of approximately 1% of the student population to almost 6% (Fig. 2(b)). In the temporal profiles, at very low adherence levels, we observed the outbreak quickly spread through the student population, peaking long before the end of term. Increasing the adherence resulted in slower spread, so that the peak occurred closer to the end of term and increasing the end of term prevalence (Fig. S8). For adherence levels above 0.7, the outbreak no longer peaked before the end of term, hence the prevalence at the end of term began to fall again, reaching a median estimate of approximately 4% (Fig. 2(b)). For all adherence levels, the majority of students infected at the end of term were non-symptomatic (either asymptomatic or presymptomatic). This was more pronounced for higher levels of infection at the end of term (Fig. 2(c)).

The level of adherence also affected the resources expended due to the isolation, test and trace interventions. We consider the amount of time spent in isolation by students to be a significant component within this (Fig. 3(a), Table S3). An increase from low to medium adherence levels (0 - 0.5) resulted in an increase in the time spent in isolation by adhering students. However, further increases in adherence (0.5 - 1) had the opposite effect. Across all levels of adherence and averaging across the student population, adhering students were expected to spend between approximately 5% - 30% of their time in isolation throughout the course of the term. Those students resident on-campus were expected to spend a greater proportion of their time in isolation (up to 40%) compared to those off-campus (up to 25%).

**Fig. 3:**
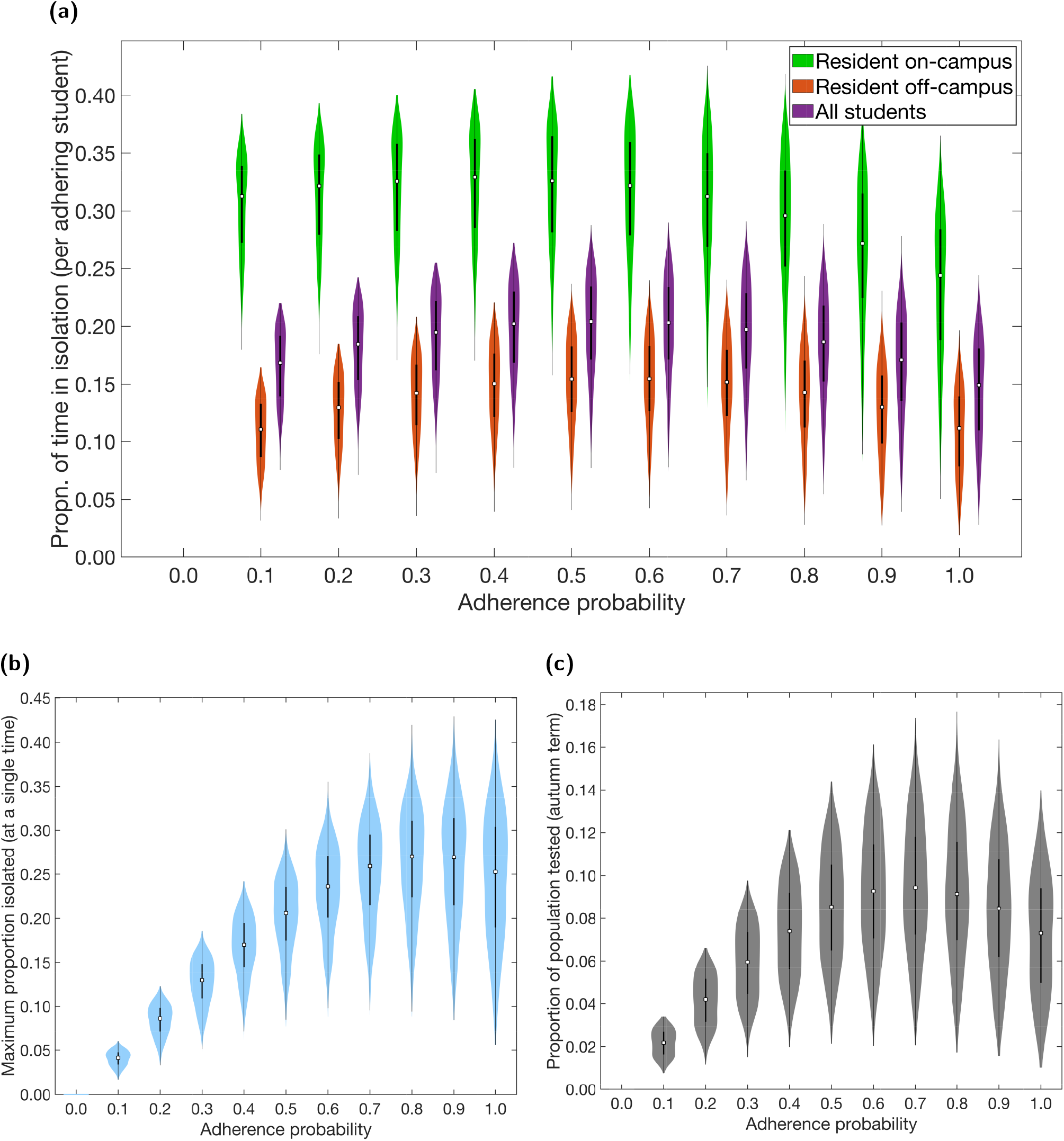
Non-infection epidemiological measures over the autumn term under differing levels of adherence to NPIs. Outputs summarised from 1,000 simulations (with 20 runs per network, for 50 network realisations) for various levels of adherence to NPIs. In all the violin plots, the white markers denote medians and solid black lines span the 25th to 75th percentiles. **(a)** Over the duration of the autumn term, distributions relative to students resident on-campus only (green violin plots), students resident off-campus only (orange violin plots) and to the overall student population (purple violin plots) for proportion of time adhering students spend in isolation. Maintenance of nonpharmaceutical interventions and effective contact tracing reduced the expected time an adhering student would spend in isolation. On-campus residents were more likely spend a greater proportion of time in isolation compared to students living off-campus. For percentile summary statistics, see Table S3. **(b)** Maximum proportion of students isolated on a single day. **(c)** Proportion of population infected by SARS-CoV-2 and tested during the autumn term.

We observed a similar relationship for both the maximum proportion of students isolating at any one time and the proportion of the population tested over the term, initially increasing as adherence increased, then falling again as adherence increased further towards 1 (Figs. 3(b) and 3(c)). The maximum proportion of students isolating at any one time was highest for adherence values between 0.8 - 0.9, on average peaking at between 20% - 30% of the student population (Fig. 3(b)). The median estimate for the proportion of students tested during the term reached a maximum of almost 10%, at adherence levels close to 0.7 (Fig. 3(c)).

The non-monotonic relationship between adherence and resource expenditure is due to the interplay between the number of students isolating and infections levels: if more students adhere, then more students will get tested and isolate when necessary, increasing the number of tests and possibly the time and number of students in isolation. However, if the increased testing and isolation of students causes a large enough reduction in prevalence over the term, this will lead to an overall reduction in the expenditure of both resources. This trade-off between isolation and prevalence can be seen in the temporal profiles of infection prevalence and proportion of students isolating (Fig. S8).

We note that, for a fixed adherence, there was significant variation in infection and resource expenditure outcomes. This was primarily due to the variability in epidemiological factors between simulation runs, such as the distribution of initial infections, the asymptomatic probability, and the relative infectiousness of an asymptomatic case, all of which were randomly generated at the start of each simulation. The different network structures contributed relatively less variation. Results from a collection of simulations performed on a single network realisation also display a large amount of variation (Figs. S9,S10).

### Use of room isolation

Room isolation for symptomatic students has been proposed as an additional measure of social distancing and control. Overall, across all adherence levels, the added use of room isolation had only a marginal impact on all previously reported measures related to both infection levels and resource expenditure (Fig. 4).

**Fig. 4:**
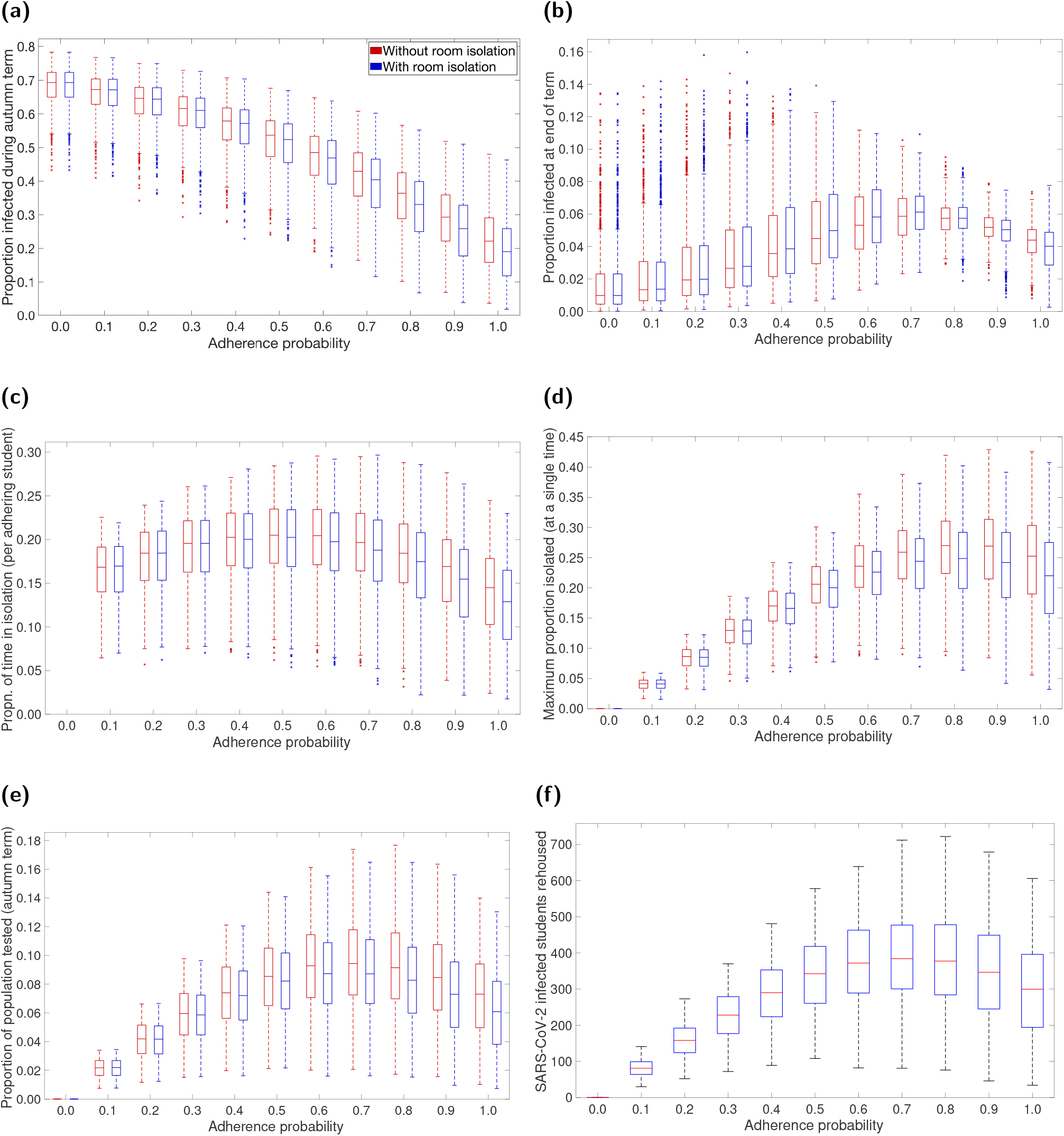
Impact on epidemiological measures across the student population of including rehousing/room isolation as part of the intervention strategy. For specified adherence levels, we compare two scenarios: one without rehousing/room isolation as part of the management strategy (red boxplots), and one including rehousing/room isolation as part of the management strategy (blue boxplots). We ran 1,000 replicates for each scenario. **(a)** Maximum number of students rehoused at any one time for the additional isolation strategy. **(b)** Proportion infected. **(c)** Proportion of population infected by SARS-CoV-2 and tested. **(d)** Proportion of time adhering students spend in isolation. **(e)** Maximum proportion of students isolated at any single time. **(f)** Proportion of students infected at the end of the autumn term. For percentile summary statistics, see Table S4. The addition of a rehousing/room isolation control measure generally resulted in slight reductions in central estimates and a narrowing of distributional ranges across all measures.

We find that room isolation resulted in a small decrease in the total infections throughout the term, with most pronounced effects at higher levels of adherence (Fig. 4(a)). We also observe slightly higher levels of infection at the end of term for mid-range adherence values (Fig. 4(b)), but the opposite for adherence values close to 1 (once more due to the interplay between the number of students isolating and infections levels).

Adding room isolation also slightly decreased the time spent in isolation and the number of tests required across all adherence levels (Figs. 4(c) to 4(e)). Room isolation also resulted in an additional outlay of resources in the form of rehousing infected students where necessary (if they did not have access to a private bathroom). Median estimates for the total number of students rehoused ranged from 0 to 400, depending on adherence levels. We observed a similar non-monotonic relationship between adherence and the number of students rehoused as previously noted for other measures of resource expenditure. The number of students rehoused was highest for adherence levels between 0.6 and 0.8 (Fig. 4(f)).

### Mass testing

We assessed the effect of mass testing on the total proportion of students infected and the proportion of time spent in isolation by adhering students. We did this for a range of mass testing scenarios, varying the timing and frequency of tests, the coverage amongst the eligible student population, and the underlying level of adherence to isolation, test and trace. Each scenario was compared to an identical scenario with mass testing removed. We report relative median values for ease of comparison between testing scenarios.

In general, implementing mass testing reduced the overall proportion of students infected throughout the term. This reduction was more pronounced at higher adherence levels, with more frequent testing and at a greater coverage of the student population. If testing could occur only once, a test date earlier in the term (end of week 2) appeared slightly more effective in reducing infections than later (end of week 8). Similarly, if only a subset of the population was to be tested, the larger amount of off-campus residents (than on-campus residents) meant testing off-campus residents only was more effective at reducing infections than testing on-campus residents only. From the scenarios considered, we found a greatest reduction in median infection levels of approximately 87% for a weekly mass testing strategy involving all students, with 80% of students adherent (Fig. 5).

**Fig. 5:**
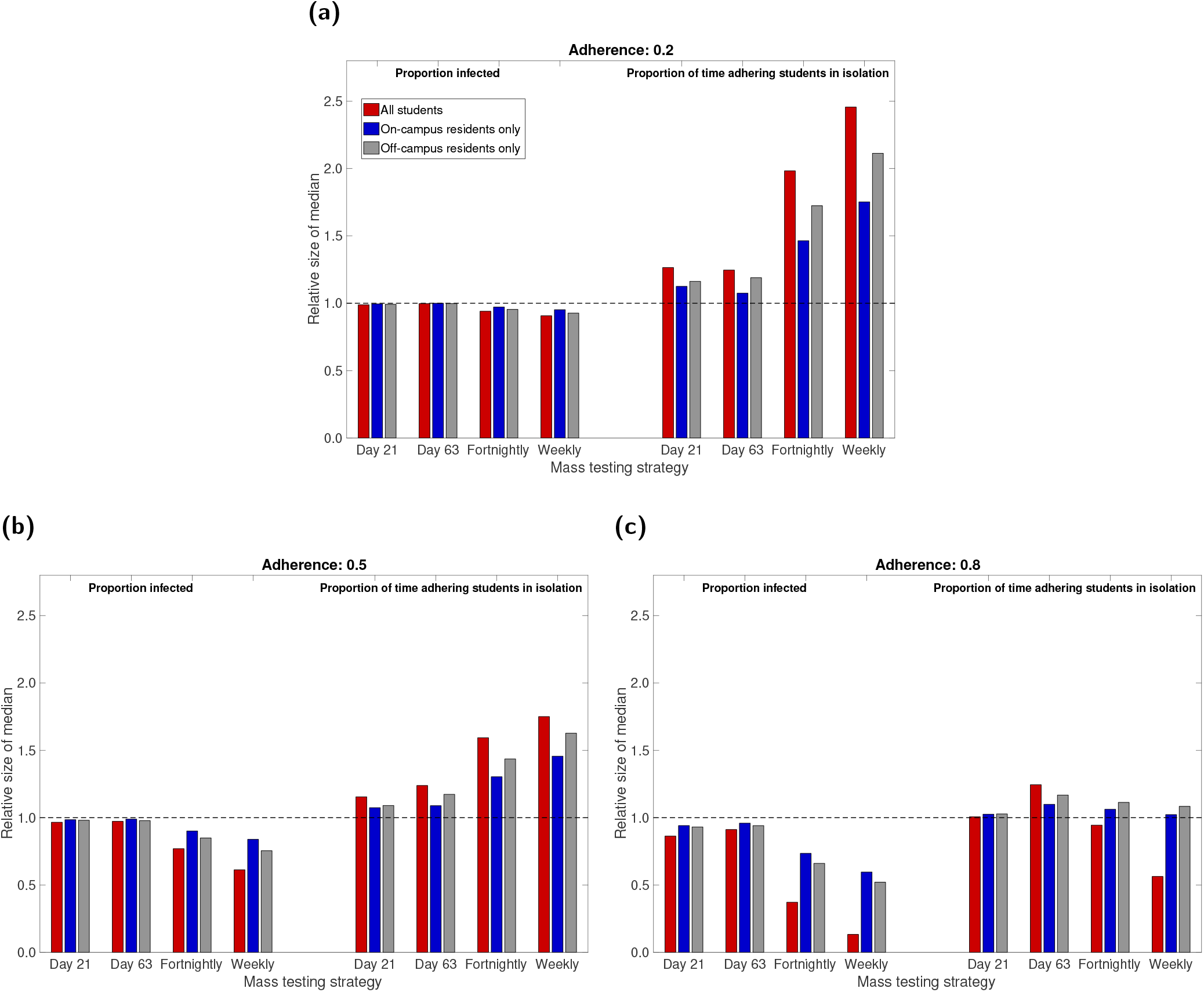
Measures of relative case load and isolation burden under the considered mass testing options. Mass testing was either not used (baseline scenario), a single instance took place at the end of week two or week eight of the academic term (corresponding to simulation day numbers 21 and 63, respectively), or regular mass testing was performed on a fortnightly or weekly basis. We present in each panel evaluations under adherence probabilities of: **(a)** 0.2 (low); **(b)** 0.5 (moderate); **(c)** 0.8 (high). Estimates in each scenario were made from 1000 simulations for mass testing covering all eligible students (red), on-campus only (blue), off-campus only (grey). The left hand side of each panel corresponds to the relative proportion (compared to the baseline scenario) of the student population infected over the duration of the autumn academic term under each mass testing strategy. In a similar way, the right hand side of each panel presents data on the relative time adherent students spent in isolation. Full estimates are given in Table S5.

The effect of mass testing on infection levels was also observed in the end of term prevalence. As previously explained, the suppression of growth can delay the peak of the outbreak and thus result in a higher prevalence at the end of term (Figs. S11-S13). We observed this for many of the mass testing scenarios considered (Fig. S14, Tables S6&S7). However, with 80% underlying adherence, regular mass testing (at any considered coverage) sufficiently suppressed the outbreak such that prevalence levels were lower at the end of term than without mass testing. This was most pronounced when all students were tested. Similarly, a one-off test at the end of week 8 also had this effect, at any level of adherence.

Mass testing generally increased the proportion of time adhering students spent in isolation, in all but two of the considered scenarios. This increase was highly dependent on the underlying adherence level, with higher levels of adherence resulting in a smaller increase. For the highest adherence level considered (0.8; Fig. 5(c)), regular testing of the entire student population led to a decrease in the time spent in isolation, due to a significantly suppressed prevalence of infection throughout the term. For lower adherence levels (Figs. 5(a) and 5(b)), increased testing frequency and greater coverage resulted in a relatively larger increase in isolation time caused by mass testing. This again displays the non-monotonic relationship between the number of students isolating and infection levels. At worst, weekly mass testing of the entire student population increased the time spent in isolation by a factor of nearly 2.5 if only 20% of students were adhering (Fig. 5(a)). However, if 80% of students adhered, the same weekly testing gave a relative median of 0.56 for time spent in isolation per adherent student, corresponding to a reduction in the time spent in isolation by approximately 45% (Fig. 5(c)).

## Discussion

In this paper, we have described the construction and application of a network model to characterise the transmission of SARS-CoV-2 amongst a student population in a UK campus-based university. Our findings suggest SARS-CoV-2 could readily transmit amongst a student population within a university setting over the course of a single academic term. Maintaining nonpharmaceutical interventions and effective contact tracing curbed transmission, however NPIs can also increase the expected time an adhering student would spend in isolation compared to without such interventions.

Our findings demonstrate the efficacy of isolation and tracing measures in controlling the spread of SARS-CoV-2 if they are broadly adhered to. Our network model results add to similar conclusions drawn from analyses using compartmental models showing that the spread of SARS-CoV-2 within a university student population can be curbed by effective testing, isolation, contract tracing and quarantine [16, 18, 35]. This is also in line with observations outside of a university setting, with the use of nonpharmaceutical interventions to control spread of SARS-CoV-2 at a national scale previously documented [36–38].

During the autumn academic term, some students were asked to complete surveys regarding their adherence to isolation and tracing measures. This work was completed prior to such data becoming available, however analysis of these surveys now offers us further insights. These empirical data indicate the majority of students reported adhering to interventions. In particular, the Student Covid Insights Survey, conducted by the Office for National Statistics, collected information in three pilots during October and November that indicated student adherence to isolation, test and trace guidance was in the region of 80-90% [39]. For example, when students were asked what actions they would take if they developed symptoms of COVID-19, between 85% and 89% of students reported that they would request a test and between 82% and 86% reported that they would stay home for 10 to 14 days (7 to 14 days in Pilot 1). When students were asked if they would share details of people that they had most recently been in contact with, if contacted by the contact tracing service, around 85% said they would be likely, or extremely likely to do so. These data suggest that our findings for higher levels of adherence are more relevant in retrospect.

Irrespective of adherence probability, we predicted that a higher proportion of the on-campus population would typically be infected compared to those living off-campus. In general, household sizes within on-campus halls of residence are larger than those living in households off-campus. As a consequence, a higher level of mixing is expected in on-campus residences, with an associated increased risk of infection. Halls of residence have been identified as environments conducive to the transmission of other respiratory illnesses [40]. This outcome reinforces the importance of monitoring the situation in halls of residence, in agreement with previous modelling work using [18].

We also analysed the impact of separating on-campus residents who were confirmed infected from household members (for the duration of the infected individual’s isolation period) could have on disease spread. We saw only marginal improvements compared to not including the intervention, and the practicalities of such a strategy and the outlay on required resources may prohibit it as an implementable option. In particular, it requires sufficient spare housing capacity with suitable facilities to accommodate those that require rehousing, as well as a safe way of moving infectious individuals to their new rooms.

Mass testing had the ability to significantly reduce overall infection levels, however only if performed regularly and when adhered to. We found that running a single mass test on the student population had only a small impact on overall infection levels. However, it also illustrated that the optimal timing of such a test was dependent on the objective. Based on our modelling framework, if one was looking to minimise the total proportion of students infected, then earlier testing would be selected. However, an additional concern is the potential risks of asymptomatic students returning home for the winter break and unwittingly spreading infection to their domiciled community. To minimise the prevalence of infection at the end of term, performing the mass test later in the term would be preferable.

Our findings regarding mass testing strategies are in agreement with prior modelling work indicating that mass testing of students would need to take place at regular intervals, such as fortnightly or weekly, to suppress SARS-CoV-2 transmission [13, 18]. There have been calls that, before universities allow students to return home, community transmission must first be curbed and frequent testing subsequently provided [41]. As an additional aid to help track and monitor the spread of COVID-19 in their student and staff communities, several universities have set up public-facing data dashboards in both the USA [42] and the UK [43].

Where possible, we have taken a data-driven approach to parameterise the system and instruct heterogeneities we expect to be present, such as in student contact patterns. Nevertheless, this work has made simplifying assumptions and our results therefore have limitations. Student numbers and estimates of regional movements between term-time and out-of-term time addresses were taken from pre-pandemic academic years; these movements may not accurately reflect the situation for the 2020/2021 academic year during the COVID-19 pandemic. Additionally, we assumed there would be no students beginning term with COVID-like symptoms, no transmission in face-to-face teaching settings and no transmission to students from the wider community. These assumptions may lead to an underestimate of transmission potential, with relaxation of any of these assumptions likely to generate a larger outbreak throughout the term.

We assumed, for simplicity, that each student maintained consistent and unchanging household, study and social contacts throughout the entire term. While the assumption for households may reasonably hold, given shared use of communal spaces, one would expect less rigidity in the study and organised social group related contacts. We also used a fixed distribution for drawing random daily social contacts throughout the term, whereas in reality we might expect the distribution of such contacts to vary temporally. A set of distributions could instead be used to capture these temporal heterogeneities, were the necessary data available to determine the distinct time periods and parameterise each distribution. Finally, the level of transmission through this network is contingent on the behaviour of students and their compliance with social distancing measures. We have assumed an uncontrolled reproduction number in the range 2-5 (dependent on the proportion of students that are asymptomatic and the relative transmission rate from asymptomatic infections); unfortunately, the precise value can only be estimated once students return and any emerging outbreak can be measured. In the event of student populations at universities suffering outbreaks, there is scope for the network model framework presented here to be used for real-time parameter estimation. Larger values of *R* are likely to result in a higher number of cases and greater pressure being exerted on test and trace services earlier in the term.

Multiple refinements of the model structure are still possible and may yield a better understanding of the outbreak impact on the broader university community. We have not included university staff members, or infection to and from the local community. Students with asymptomatic infection interacting with elder individuals in non-COVID secure environments may result in silent transmission of SARS-CoV-2 into more vulnerable groups at risk of severe outcomes from COVID-19. Similarly, given that older individuals are at greater risk of severe health outcomes due to COVID-19 disease [7], in the event of widespread community transmission, staff and surrounding communities would be likely to experience higher levels of morbidity than students. Another aspect we have not included here is the presence of other respiratory infections. Such an extension would permit the study of test capacity requirements when levels of cough and fever are high due to non-COVID-19 causes, especially of concern in the winter period; were such a scenario to arise it would apply significant stress to the national test and trace system [44].

In the context of the COVID-19 pandemic the movement of students to attend universities, creating large communities of predominately young adults, poses specific challenges in controlling transmission. Infectious disease models may be a useful part of the public health decision-making process, determining the most appropriate interventions to be applied in a university setting. Our work highlights a network modelling approach to capture heterogeneities in contact structure that are particular to the university student population and its projected impact on transmission of SARS-CoV-2. This model suggests that encouraging student adherence with test-trace-and-isolate rules (as well as good social-distancing, mask-use and hygiene practices) is likely to lead to the greatest reduction in cases both during and at the end of term; mass testing is also found to produce strong benefits in terms of reducing infection, generally leading to a greater number of cases being found and isolated.

## Data Availability

Summary data that were used to perform the study are publicly available or stated within the main manuscript and Supporting Information. Code for the study is available at https://github.com/EdMHill/covid19_uni_network_model.

## Author contributions

**Edward M. Hill:** Conceptualisation, Data curation, Formal analysis, Methodology, Software, Validation, Visualisation, Writing - Original Draft, Writing - Review & Editing.

**Benjamin D. Atkins:** Conceptualisation, Data curation, Formal analysis, Methodology, Software, Validation, Visualisation, Writing - Original Draft, Writing - Review & Editing.

**Matt J. Keeling:** Conceptualisation, Data curation, Funding acquisition, Methodology, Writing - Review & Editing.

**Michael J. Tildesley:** Conceptualisation, Data curation, Funding acquisition, Methodology, Software, Supervision, Validation, Writing - Review & Editing.

**Louise Dyson:** Conceptualisation, Data curation, Funding acquisition, Methodology, Software, Supervision, Validation, Writing - Review & Editing.

