## Supporting Information for "Modelling SARS-CoV-2 transmission in a UK university setting"

#### Table of Contents

**1 Supporting Text S1: Off-campus student household size** **2**

**2 Supporting Text S2: Cohort data** **3**

**3 Supporting Text S3: Parameterisation of contact risk** **6**

**4 Supporting Text S4: Non-intervention scenario calibration** **7**

**5 Supporting Text S5: Initial seeding of infected and recovered individuals** **10**

**6 Additional tables** **11**

**7 Additional figures** **13**

### 1 Supporting Text S1: Off-campus student household size

To gauge the typical sizes of off-campus student households, we used data from the Social Contact Survey [1–3], a paper-based and online survey of 5,388 participants in Great Britain conducted in 2010.

We extracted records provided by 347 students. For these participants, we isolated contacts recorded as occurring in the home setting and having a frequency in the highest category. We then increased all numbers by one to obtain household sizes including the respondent. Respondents that reported no daily household contacts we assumed to be in a household size of one. To generate the off-campus households, we fit a lognormal distribution to the data (whose best-fit parameters were mean 0.979 and standard deviation 0.576) and sampled the size of each household from the associated probability density function (Fig. S1). We subsequently assigned the required number of students to that household.

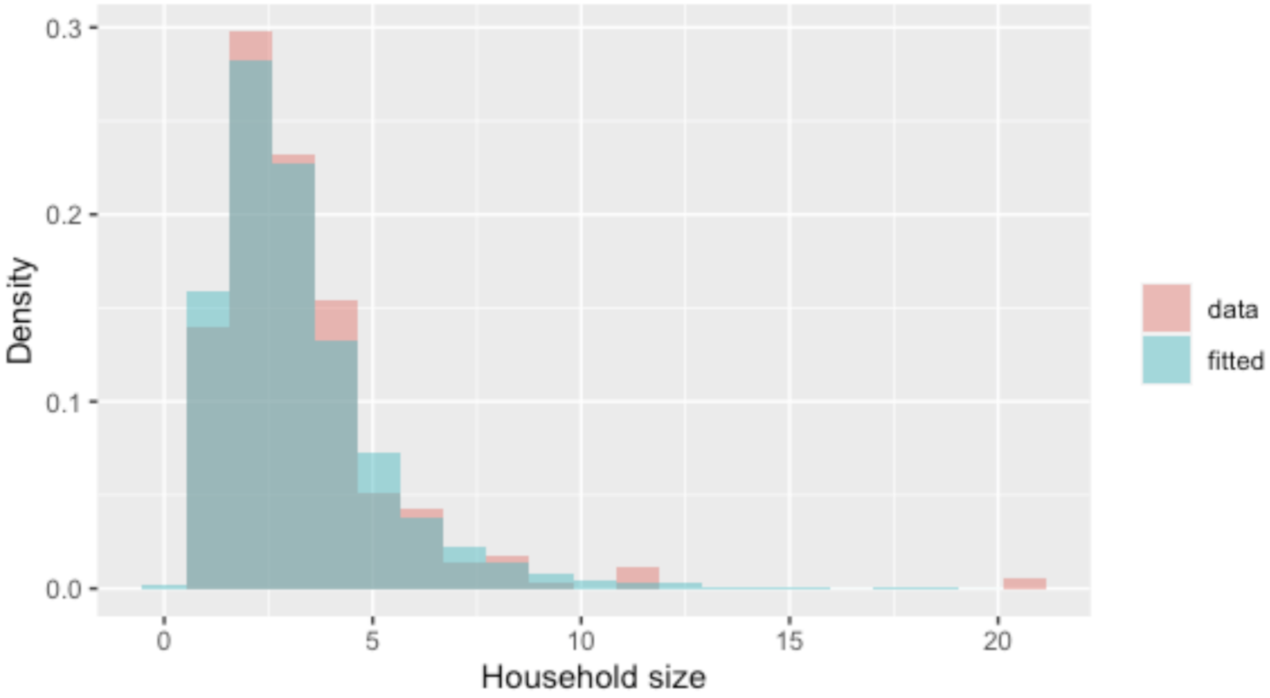


**Fig. S1: Off-campus household size probability mass functions.** Red bars represent the empirical distribution. Blue bars show the probability density function produced by the lognormal(0.979,0.576) distribution. To generate the off-campus households in our analysis, we used the fitted lognormal distribution.

#### 2 Supporting Text S2: Cohort data

We were provided with records of on-campus resident students for the 2019/2020 academic year from a representative campus based university. Each student record had information on the department of study and whether the student was in undergraduate or postgraduate study.

For computing the departmental distributions for those resident off-campus, we used high level faculty proportion statistics for the 2018/2019 academic year to obtain the relative proportion of off-campus residents within each faculty (i.e. accounting for the portions of each faculty population attributed to on-campus residents). To simplify the calculation, we assumed all first-year undergraduates were either resident on-campus or were distance learners and therefore not living in the local vicinity of the university. We evaluated department specific allocations using the same fractions as given by the data for on-campus residents.

In all, we retained 28 department labels. We calculated separate departmental breakdowns for those resident on-campus (Fig. S2(a) and Table S1) and off-campus (Fig. S2(b) and Table S2) from the available data.

**Table S1:** On-campus residence students stratified by department and study type (first year undergraduate, non-first year undergraduate or postgraduate). Percentages are given to 2.d.p.

| Faculty | Label | Department | First year undergraduate | Non-first year undergraduate | Postgraduate |
|---|---|---|---|---|---|
| **Arts** | LN | Modern Languages | 2.44 | 1.23 | 0.12 |
| | HA | History of Art | 0.53 | 0.04 | 0.00 |
| | HI | History | 4.13 | 0.26 | 0.03 |
| | TH | Theatre Studies | 0.28 | 0.01 | 0.26 |
| | EN | English | 2.50 | 0.18 | 0.22 |
| | GD | Global development | 1.69 | 0.35 | 0.00 |
| | CX | Classics | 0.72 | 0.09 | 0.01 |
| | FI | Film and Lit. | 0.85 | 0.03 | 0.03 |
| | IP | Liberal Arts | 0.46 | 0.00 | 0.00 |
| **Social Studies** | IB | Business School | 11.62 | 0.51 | 5.58 |
| | EC | Economics | 6.42 | 0.21 | 1.01 |
| | PO | Politics | 3.61 | 0.29 | 0.50 |
| | PH | Philosophy | 4.76 | 0.22 | 0.13 |
| | LA | Law | 3.63 | 0.21 | 0.31 |
| | SO | Sociology | 1.15 | 0.16 | 0.21 |
| | ET | Applied Linguistics | 0.84 | 0.06 | 0.54 |
| | FP | Foundation Programme | 0.62 | 0.00 | 0.00 |
| | IM | Interdisciplinary Methods | 0.00 | 0.00 | 0.43 |
| **Science & Engineering** | LF | Life Sciences | 4.67 | 0.35 | 0.34 |
| | MA | Mathematics | 4.89 | 0.37 | 0.16 |
| | CS | Computer Science | 4.45 | 0.21 | 0.46 |
| | ST | Statistics | 4.03 | 0.13 | 0.29 |
| | WM | Manufacturing | 0.75 | 0.01 | 4.82 |
| | ES | Engineering | 5.07 | 0.32 | 0.26 |
| | PS | Psychology | 2.67 | 0.18 | 0.25 |
| | PX | Physics | 3.16 | 0.13 | 0.03 |
| | CH | Chemistry | 2.13 | 0.26 | 0.09 |

**Table S2:** Off-campus residence students stratified by department and study type (first year undergraduate, non-first year undergraduate or postgraduate). Percentages are given to 2.d.p.

| Faculty | Department | First year undergraduate | Non-first year undergraduate | Postgraduate |
|---|---|---|---|---|
| **Arts** | Modern Languages | 0 | 2.02 | 0.40 |
| | History of Art | 0 | 0.31 | 0.00 |
| | History | 0 | 2.41 | 0.10 |
| | Theatre Studies | 0 | 0.16 | 0.89 |
| | English | 0 | 1.47 | 0.74 |
| | Global development | 0 | 1.12 | 0.00 |
| | Classics | 0 | 0.44 | 0.05 |
| | Film and Lit. | 0 | 0.48 | 0.10 |
| | Liberal Arts | 0 | 0.25 | 0.00 |
| **Social Studies** | Business School | 0 | 7.23 | 15.83 |
| | Economics | 0 | 3.95 | 2.87 |
| | Politics | 0 | 2.33 | 1.42 |
| | Philosophy | 0 | 2.97 | 0.37 |
| | Law | 0 | 2.28 | 0.87 |
| | Sociology | 0 | 0.78 | 0.58 |
| | Applied Linguistics | 0 | 0.53 | 1.54 |
| | Foundation Programme | 0 | 0.37 | 0.00 |
| | Interdisciplinary Methods | 0 | 0.00 | 1.21 |
| **Science & Engineering** | Life Sciences | 0 | 3.05 | 0.79 |
| | Mathematics | 0 | 3.20 | 0.38 |
| | Computer Science | 0 | 2.83 | 1.06 |
| | Statistics | 0 | 2.53 | 0.68 |
| | Manufacturing | 0 | 0.46 | 11.23 |
| | Engineering | 0 | 3.28 | 0.62 |
| | Psychology | 0 | 1.73 | 0.58 |
| | Physics | 0 | 2.00 | 0.07 |
| | Chemistry | 0 | 1.45 | 0.21 |
| **Medicine** | All medical school | 0 | 4.19 | 3.57 |

**(a)**

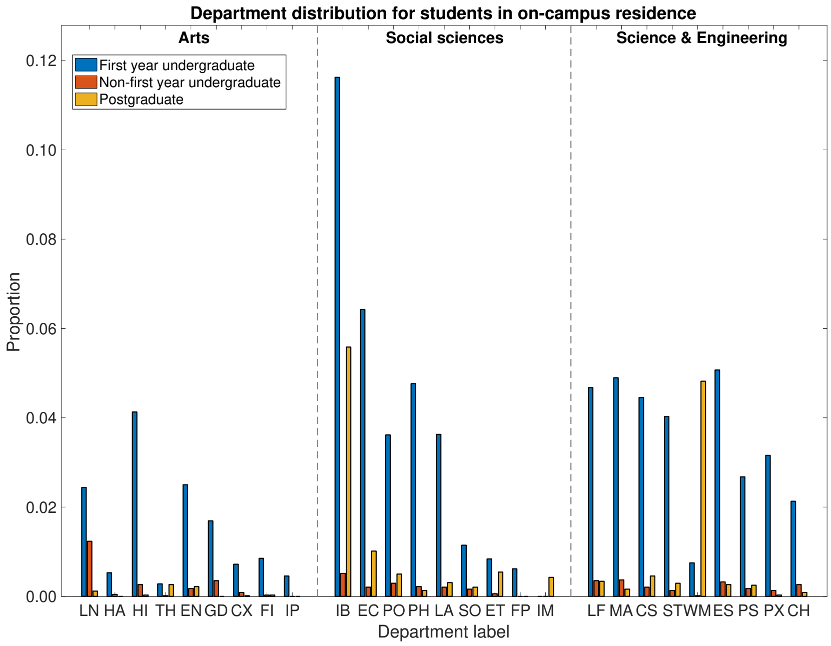


**(b)**

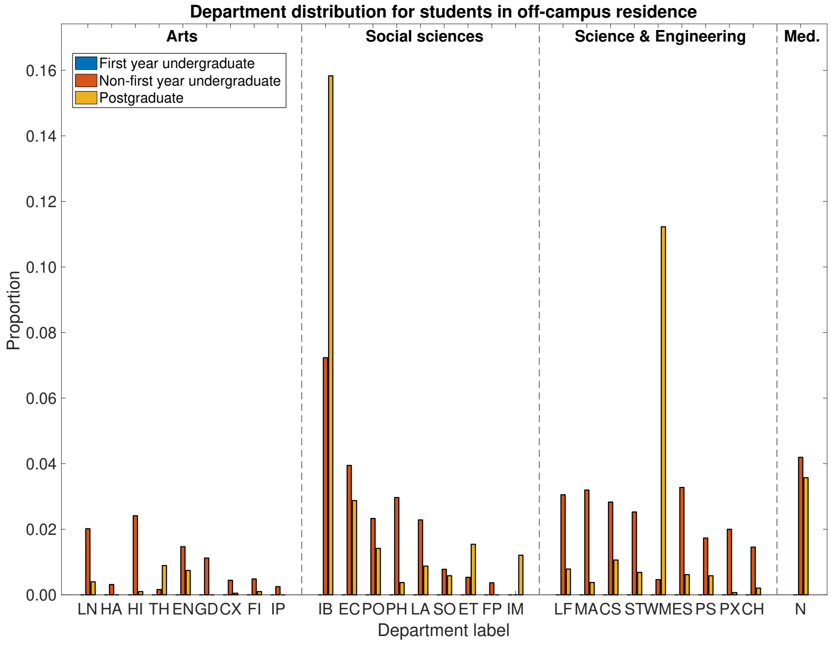


**Fig. S2: Per residence location, the proportion of students within each department.** Panels correspond to: **(a)** Students resident on-campus; **(b)** Students living off-campus. In each panel, blue bars represent first year undergraduate students, orange bars represent non-first year undergraduate students and yellow bars represent postgraduate students. We list department labels in Table S1.

### 3 Supporting Text S3: Parameterisation of contact risk

The data contained in the Social Contact Survey [1–3] included participants recording for each interaction its duration and whether it involved physical touch. We used these data attributes to scale the transmission risk of contacts occurring in non-household settings relative to household contacts.

Explicitly, the contact survey found 80% of household contacts to involve touch. As a result, touch contacts contributed 80% to the household secondary attack rate estimate. The remaining 20% of the household secondary attack rate we attributed to non-touch contacts with a duration classification above 0; of the non-touch contacts, 80% had duration classified above 0.

For contacts in non-household settings, relative to the central estimate of adjusted secondary attack rate in the household setting against those aged 18-34 of 0.34 [4], the above procedure returned a transmission risk value of 0.2414. The relative magnitude of that estimate was used to scale the standard deviation, consequently set at 0.034. We applied these computed transmission risks for contacts in the study (cohort) setting, dynamic social contacts and within sports clubs.

For societies, we assigned a lower transmission risk to reflect the implementation of COVID-secure measures that would be required to permit these meetings to take place (mean 0.12, standard deviation 0.017).

For each individual, we drew the transmission potential across contacts in each respective setting from normal distributions with the specified mean and standard deviation values.

### 4 Supporting Text S4: Non-intervention scenario calibration

#### Model parameterisation

In the absence of isolation and contact tracing, we calibrated the system to return a 7-day moving average $R_t$ in the range of three to four in the early phase of the outbreak (Fig. S3). We achieved this magnitude of spread by applying a scaling factor of 0.8 to the baseline transmission risk across a contact in each setting.

Relative to the assumption made in the main manuscript of society and sports clubs being run in a COVID-secure fashion, for the no intervention scenario considered here we consequently attributed a higher risk of transmission for contacts occurring in these organised social groups. The baseline transmission risk in societies matched that in non-household settings. In sports clubs, the baseline transmission risk matched the central estimate of adjusted secondary attack rate in the household setting against those aged 18-34 [4].

For each replicate, we drew the probability of a case being asymptomatic from a Uniform(0.5,0.8) distribution and the relative infectiousness of an asymptomatic from a Uniform(0.3,0.7) distribution. All simulations had a duration of 77 days (11 weeks). We ran batches of 100 stochastic simulations on different network configurations, as well as a run of 1,000 simulations whose realisations consisted of 50 separate networks (with 20 runs performed using each network). All individuals began susceptible, with the exception of ten individuals seeded as initial infecteds (five symptomatic, five asymptomatic).

#### Results summary

For the early stages of the outbreak, in our simulations batches we typically obtained a distribution for the 7-day moving average $R_t$ whose 50% prediction interval lay in the range 3 to 4 (for an example, see Fig. S3).

Across our simulated collection of mean generation times the median value was roughly seven days (Fig. S4). In the absence of interventions, a mean generation time estimate in the region of seven days corresponds with findings from an analysis of transmission pair data from mainland China by Ali *et al.* [5], which found serial intervals were on average 7.8 days in mid-January 2020 (prior to the implementation of nonpharmaceutical interventions).

In general, up to 95% of the student population was infected by the conclusion of autumn term. The contribution of static contacts to infection was greatest in dynamic social contacts, followed by cohort contacts and household contacts. Organised society and sports club meeting, that occur less frequently, provided a smaller contribution to overall transmission. Finally, we observed a non-negligible proportion of infection occurring due to dynamic contacts in accommodation blocks (Fig. S5).

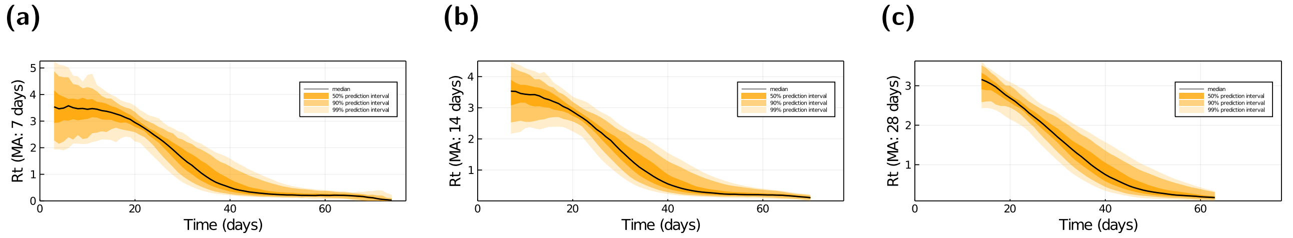


**Fig. S3: Effective reproductive ratio temporal profiles for specified centralised moving average widths.** In each panel, the solid line represents the median estimate, with shaded regions illustrating the 50%, 90% and 99% prediction intervals, respectively (transitioning from darkest to lightest). We display temporal profiles of $R_t$ with centralised moving averages of: **(a)** 7 days; **(b)** 14 days; **(c)** 28 days. Note, the y-axis scales differ across panels.

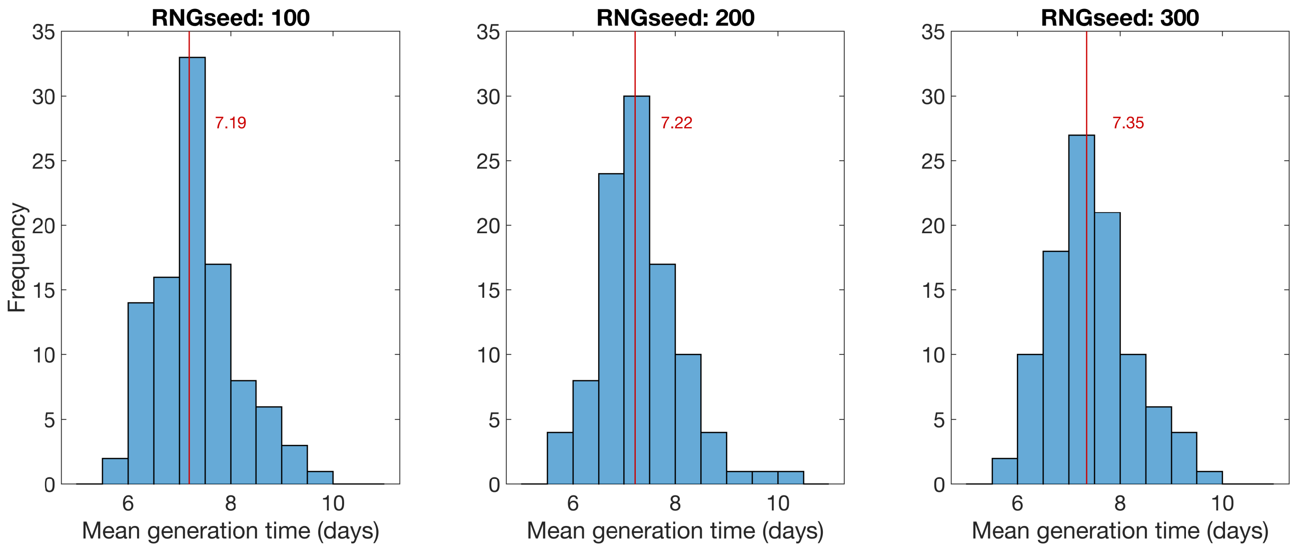


**Fig. S4: Distribution of mean generation times from 100 replicates.** Each panel was produced with the random number generator passed a different seed. The vertical line designates the median value, with the corresponding value (to 2 d.p.) stated alongside the line. We obtained a median value for the mean generation time of roughly seven days.

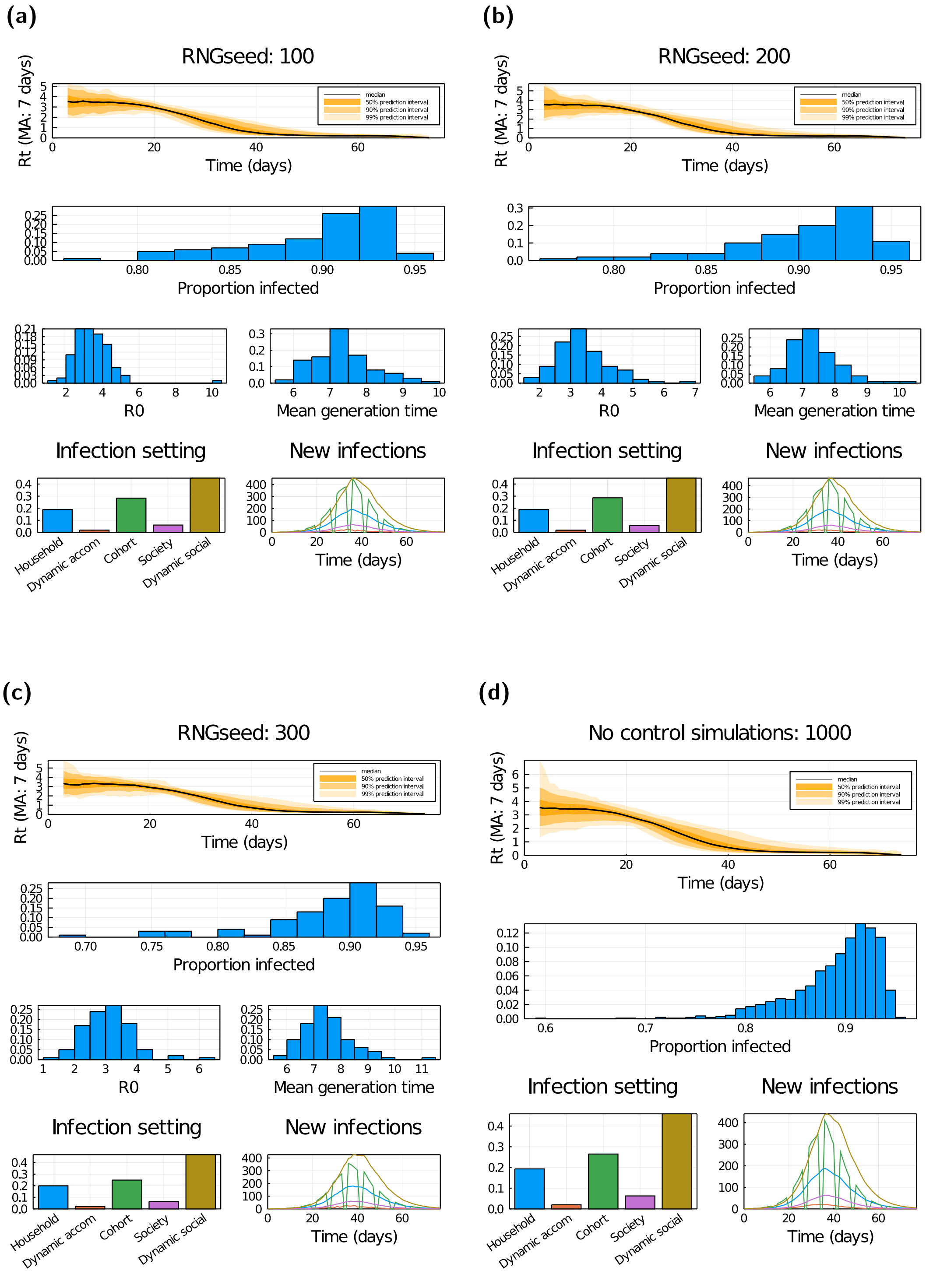


**Fig. S5: Outbreak temporal profiles and associated probability distributions in the absence of interventions. (a-c)** Estimated from three separate batches of 100 stochastic simulations, each on a different network realisation. *(Row one)* Daily $R_t$ estimate using a 7-day moving average. *(Row two)* Proportion of individuals infected after eleven weeks have elapsed. *(Row three, left)* Average number of individuals infected by the initial ten nodes. *(Row three, right)* Mean generation time. *(Row four, left)* Proportion of infections occurring in each setting. *(Row four, right)* Average (mean) amount of new daily infections in each setting. **(d)** Outputs summarised from 1,000 simulations (consisting of 50 network realisations and 20 runs per network). Panels match those presented in rows one, two and four for **(a-c)**.

### 5 Supporting Text S5: Initial seeding of infected and recovered individuals

We initialised the number of incoming infected students (stratified by latent and asymptomatic states) and incoming students that had previously been infected (assumed recovered) by multiplying the number of incoming students to the university from a UK region by the percentage of the incoming student population from that region estimated to be of that COVID-19 disease status (distribution of sampled values shown in Fig. S6). For each individual initialised in the latent infected or asymptomatic disease state, we also sampled the time they had already spent in the respective disease state.

To inform the student flows, we used rounded figures provided from HESA via Jisc on the number of students at each HE provider who are domiciled in the different English regions and the devolved administrations. These numbers were from the 2019/2020 academic year, and excluded distance-learning students and students on industrial placements (https://github.com/magicicada/simple_epi_calculations/tree/main/basic_arriving_student_calculations).

For estimating the proportion of the student population that resided in each disease compartment upon commencement of the university term, we drew on projections from a pre-existing SARS-CoV-2 ODE transmission model [6]. Values were taken from 26th September, corresponding with the beginning of welcome week.

We assumed no students arrived with symptomatic infection. In addition, we only accounted for infection arriving from students domiciled in the UK. We make a simplifying assumption that non-UK students would not contribute to the initial infected count, with an argument that argue that the majority of international students would be required to quarantine for 14 days after entering the UK. However, we did apply a generalising assumption of the proportion of international-based students initialised in the recovered state to match the estimated proportion of arriving UK-based students in the recovered state.

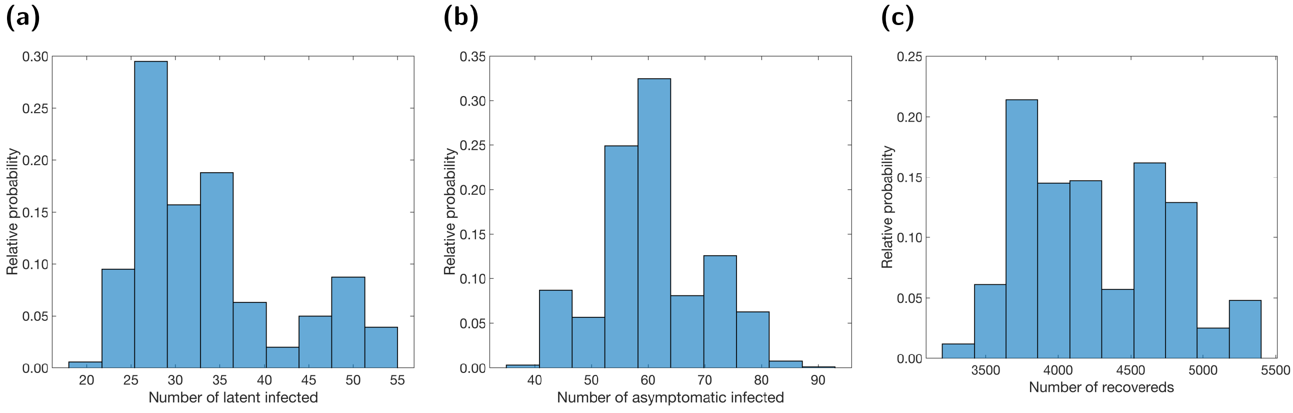


**Fig. S6: Distributions of the number of students with the given COVID-19 disease status at the beginning of welcome week.** We produced the displayed histograms from 100 simulations. **(a)** Latent infected; **(b)** asymptomatic infecteds; **(c)** previously infected (assumed recovered).

### 6 Additional tables

**Table S3:** Summary statistics for proportion infected and proportion of time adhering individuals spend in isolation dependent upon both the residence location and the probability an individual adheres to intervention measures. We present median estimates and give 95% prediction intervals in parentheses, produced from 1000 simulation replicates. All values given to 2 s.f. − − − corresponds to a scenario where the specified metric is not relevant.

| | Proportion adhering | | | | | | | | | | |
|---|---|---|---|---|---|---|---|---|---|---|---|
| **Proportion infected** | **0.0** | **0.1** | **0.2** | **0.3** | **0.4** | **0.5** | **0.6** | **0.7** | **0.8** | **0.9** | **1.0** |
| **On-campus** | 0.77 (0.67,0.82) | 0.75 (0.64,0.80) | 0.73 (0.61,0.79) | 0.71 (0.57,0.77) | 0.68 (0.53,0.75) | 0.65 (0.47,0.73) | 0.62 (0.42,0.71) | 0.57 (0.36,0.68) | 0.52 (0.29,0.65) | 0.45 (0.21,0.61) | 0.37 (0.14,0.57) |
| **Off-campus** | 0.66 (0.51,0.74) | 0.64 (0.48,0.72) | 0.61 (0.44,0.69) | 0.58 (0.40,0.66) | 0.54 (0.34,0.63) | 0.49 (0.28,0.60) | 0.43 (0.23,0.55) | 0.37 (0.18,0.51) | 0.30 (0.13,0.46) | 0.23 (0.085,0.41) | 0.16 (0.048,0.35) |
| **All** | 0.69 (0.56,0.76) | 0.67 (0.53,0.74) | 0.65 (0.49,0.72) | 0.62 (0.45,0.70) | 0.58 (0.40,0.67) | 0.54 (0.34,0.64) | 0.49 (0.29,0.60) | 0.43 (0.23,0.56) | 0.36 (0.18,0.51) | 0.29 (0.12,0.46) | 0.22 (0.076,0.41) |
| **Proportion of time isolated** | | | | | | | | | | | |
| **On-campus** | — | 0.31 (0.22,0.36) | 0.32 (0.22,0.37) | 0.33 (0.22,0.38) | 0.33 (0.21,0.39) | 0.33 (0.20,0.39) | 0.32 (0.19,0.39) | 0.31 (0.17,0.39) | 0.30 (0.15,0.38) | 0.27 (0.13,0.36) | 0.24 (0.10,0.33) |
| **Off-campus** | — | 0.11 (0.056,0.15) | 0.13 (0.067,0.17) | 0.14 (0.074,0.19) | 0.15 (0.078,0.20) | 0.15 (0.075,0.21) | 0.15 (0.073,0.22) | 0.15 (0.065,0.21) | 0.14 (0.06,0.20) | 0.13 (0.05,0.19) | 0.11 (0.038,0.17) |
| **All** | — | 0.17 (0.10,0.21) | 0.18 (0.11,0.23) | 0.19 (0.12,0.24) | 0.20 (0.12,0.26) | 0.20 (0.11,0.26) | 0.20 (0.11,0.27) | 0.20 (0.096,0.26) | 0.19 (0.086,0.25) | 0.17 (0.074,0.24) | 0.15 (0.057,0.22) |

**Table S4:** Summary statistics under various isolation adherence assumptions. We present median estimates and give 95% prediction intervals in parentheses, produced from 1000 simulation replicates. All values given to 2 s.f. − − − corresponds to a scenario where the specified metric is not relevant.

| | Proportion adhering | | | | | | | | | | |
|---|---|---|---|---|---|---|---|---|---|---|---|
| **No room isolation** | **0.0** | **0.1** | **0.2** | **0.3** | **0.4** | **0.5** | **0.6** | **0.7** | **0.8** | **0.9** | **1.0** |
| **Proportion infected** | 0.69 (0.56,0.76) | 0.67 (0.53,0.74) | 0.65 (0.49,0.72) | 0.62 (0.45,0.70) | 0.58 (0.40,0.67) | 0.54 (0.34,0.64) | 0.49 (0.29,0.60) | 0.43 (0.23,0.56) | 0.36 (0.18,0.51) | 0.29 (0.12,0.46) | 0.22 (0.076,0.41) |
| **Proportion of time isolated** | 0.00 (0.00,0.00) | 0.17 (0.10,0.21) | 0.18 (0.11,0.23) | 0.19 (0.12,0.24) | 0.20 (0.12,0.26) | 0.20 (0.11,0.26) | 0.20 (0.11,0.27) | 0.20 (0.096,0.26) | 0.19 (0.086,0.25) | 0.17 (0.074,0.24) | 0.15 (0.057,0.22) |
| **Proportion tested** | 0.00 (0.00,0.00) | 0.022 (0.011,0.032) | 0.042 (0.021,0.061) | 0.060 (0.029,0.087) | 0.074 (0.035,0.11) | 0.085 (0.039,0.13) | 0.093 (0.041,0.14) | 0.094 (0.039,0.15) | 0.091 (0.036,0.15) | 0.085 (0.031,0.14) | 0.073 (0.023,0.12) |
| **Maximum proportion isolated** | 0.00 (0.00,0.00) | 0.041 (0.023,0.055) | 0.086 (0.048,0.11) | 0.13 (0.073,0.17) | 0.17 (0.097,0.22) | 0.21 (0.12,0.27) | 0.24 (0.13,0.31) | 0.26 (0.14,0.34) | 0.27 (0.14,0.36) | 0.27 (0.13,0.37) | 0.25 (0.10,0.37) |
| **Proportion infected at end of term** | 0.0098 (0.0014,0.084) | 0.013 (0.0022,0.093) | 0.019 (0.0041,0.095) | 0.027 (0.0063,0.10) | 0.036 (0.01,0.10) | 0.045 (0.016,0.10) | 0.053 (0.022,0.097) | 0.059 (0.031,0.088) | 0.057 (0.038,0.077) | 0.052 (0.036,0.069) | 0.044 (0.019,0.062) |
| **With Room isolation** | | | | | | | | | | | |
| **Proportion infected** | 0.69 (0.56,0.76) | 0.67 (0.52,0.74) | 0.64 (0.48,0.72) | 0.61 (0.43,0.69) | 0.57 (0.37,0.66) | 0.52 (0.31,0.63) | 0.47 (0.25,0.59) | 0.40 (0.19,0.54) | 0.33 (0.13,0.50) | 0.26 (0.082,0.45) | 0.19 (0.047,0.39) |
| **Proportion of time isolated** | 0.00 (0.00,0.00) | 0.17 (0.10,0.21) | 0.18 (0.11,0.23) | 0.19 (0.12,0.24) | 0.20 (0.11,0.26) | 0.20 (0.10,0.26) | 0.20 (0.096,0.26) | 0.19 (0.087,0.26) | 0.18 (0.07,0.25) | 0.16 (0.057,0.23) | 0.13 (0.041,0.20) |
| **Proportion tested** | 0.00 (0.00,0.00) | 0.022 (0.011,0.032) | 0.042 (0.021,0.061) | 0.059 (0.029,0.086) | 0.072 (0.034,0.11) | 0.082 (0.035,0.13) | 0.087 (0.036,0.14) | 0.087 (0.033,0.14) | 0.083 (0.026,0.14) | 0.073 (0.023,0.12) | 0.061 (0.016,0.11) |
| **Maximum proportion isolated** | 0.00 (0.00,0.00) | 0.041 (0.023,0.055) | 0.085 (0.048,0.11) | 0.13 (0.072,0.17) | 0.17 (0.093,0.22) | 0.20 (0.11,0.26) | 0.23 (0.12,0.30) | 0.24 (0.12,0.33) | 0.25 (0.11,0.35) | 0.24 (0.099,0.35) | 0.22 (0.071,0.35) |
| **Proportion infected at end of term** | 0.0098 (0.0014,0.084) | 0.014 (0.0023,0.092) | 0.020 (0.0042,0.10) | 0.028 (0.0067,0.11) | 0.039 (0.011,0.11) | 0.050 (0.017,0.10) | 0.058 (0.025,0.097) | 0.061 (0.034,0.086) | 0.057 (0.037,0.078) | 0.050 (0.022,0.068) | 0.040 (0.010,0.063) |
| **Students rehoused** | 0 (0,0) | 81 (42,122) | 158 (85,232) | 228 (119,334) | 290 (144,428) | 342 (166,512) | 372 (170,573) | 384 (158,598) | 378 (138,622) | 346 (112,582) | 300 (81,519) |

**Table S5:** Measures of relative case load and isolation burden under the considered adherence probabilities and mass testing options compared to a scenario where no mass test was performed. All values are specified to 2 d.p.

| | | Relative proportion infected | | | Relative time adhering student isolated | | |
|---|---|---|---|---|---|---|---|
| **Adherence** | **Test strategy** | **All** | **On-campus** | **Off-campus** | **All** | **On-campus** | **Off-campus** |
| **Low (probability: 0.2)** | **Day 21** | 0.99 | 1.00 | 0.99 | 1.26 | 1.13 | 1.16 |
| | **Day 63** | 1.00 | 1.00 | 1.00 | 1.25 | 1.07 | 1.19 |
| | **Fortnightly** | 0.94 | 0.97 | 0.95 | 1.98 | 1.46 | 1.72 |
| | **Weekly** | 0.91 | 0.95 | 0.93 | 2.46 | 1.75 | 2.11 |
| **Moderate (probability: 0.5)** | **Day 21** | 0.97 | 0.98 | 0.98 | 1.16 | 1.07 | 1.09 |
| | **Day 63** | 0.97 | 0.99 | 0.98 | 1.24 | 1.09 | 1.17 |
| | **Fortnightly** | 0.77 | 0.90 | 0.85 | 1.59 | 1.30 | 1.44 |
| | **Weekly** | 0.61 | 0.84 | 0.76 | 1.75 | 1.46 | 1.63 |
| **High (probability: 0.8)** | **Day 21** | 0.86 | 0.94 | 0.93 | 1.01 | 1.03 | 1.03 |
| | **Day 63** | 0.91 | 0.96 | 0.94 | 1.24 | 1.10 | 1.17 |
| | **Fortnightly** | 0.37 | 0.73 | 0.66 | 0.94 | 1.06 | 1.11 |
| | **Weekly** | 0.13 | 0.60 | 0.52 | 0.56 | 1.02 | 1.08 |

**Table S6:** Median and 95% prediction intervals for end of term infection prevalence under the tested combinations of isolation, test and tracing adherence, mass test day and mass test coverage. All values are specified to 2 s.f. Fields containing − − − designate non-applicable combinations of test day and coverage.

| | | Mass testing coverage | | | |
|---|---|---|---|---|---|
| **Adherence** | **Test day** | **None** | **All** | **On-campus** | **Off-campus** |
| **0.2** | **None** | 0.020 (0.0042,0.10) | — | — | — |
| | **Day 21** | — | 0.026 (0.0064,0.11) | 0.023 (0.0050,0.11) | 0.023 (0.0057,0.11) |
| | **Day 63** | — | 0.018 (0.0038,0.088) | 0.019 (0.0038,0.095) | 0.018 (0.0039,0.090) |
| | **Fortnightly** | — | 0.030 (0.0072,0.11) | 0.026 (0.0057,0.11) | 0.026 (0.0062,0.10) |
| | **Weekly** | — | 0.035 (0.0089,0.12) | 0.029 (0.0071,0.11) | 0.029 (0.0074,0.10) |
| **0.5** | **None** | 0.050 (0.017,0.10) | — | — | — |
| | **Day 21** | — | 0.071 (0.032,0.12) | 0.059 (0.023,0.11) | 0.061 (0.027,0.11) |
| | **Day 63** | — | 0.036 (0.012,0.073) | 0.045 (0.016,0.090) | 0.039 (0.013,0.081) |
| | **Fortnightly** | — | 0.070 (0.036,0.10) | 0.064 (0.027,0.10) | 0.060 (0.028,0.094) |
| | **Weekly** | — | 0.071 (0.033,0.098) | 0.069 (0.033,0.10) | 0.060 (0.031,0.088) |
| **0.8** | **None** | 0.057 (0.037,0.078) | — | — | — |
| | **Day 21** | — | 0.077 (0.035,0.10) | 0.066 (0.039,0.088) | 0.067 (0.037,0.088) |
| | **Day 63** | — | 0.030 (0.018,0.040) | 0.045 (0.029,0.058) | 0.038 (0.025,0.051) |
| | **Fortnightly** | — | 0.032 (0.0047,0.057) | 0.055 (0.018,0.072) | 0.045 (0.020,0.063) |
| | **Weekly** | — | 0.0093 (0.00,0.050) | 0.049 (0.013,0.071) | 0.038 (0.014,0.053) |

**Table S7:** Performance of inclusion of mass testing, relative to a scenario where no mass test was performed, measured by end of term prevalence metrics. All values are specified to 2 d.p.

| | | Probability mass testing reduces end of term prevalence | | | Relative end of term prevalence | | |
|---|---|---|---|---|---|---|---|
| **Adherence** | **Test day** | **All** | **On-campus** | **Off-campus** | **All** | **On-campus** | **Off-campus** |
| **0.2** | **Day 21** | 0.03 | 0.14 | 0.09 | 1.29 | 1.15 | 1.18 |
| | **Day 63** | 0.92 | 0.73 | 0.90 | 0.90 | 0.97 | 0.92 |
| | **Fortnightly** | 0.07 | 0.13 | 0.17 | 1.52 | 1.29 | 1.31 |
| | **Weekly** | 0.04 | 0.06 | 0.11 | 1.74 | 1.48 | 1.44 |
| **0.5** | **Day 21** | 0.01 | 0.03 | 0.04 | 1.42 | 1.18 | 1.23 |
| | **Day 63** | 1.00 | 0.98 | 1.00 | 0.72 | 0.90 | 0.78 |
| | **Fortnightly** | 0.20 | 0.15 | 0.26 | 1.41 | 1.29 | 1.21 |
| | **Weekly** | 0.26 | 0.15 | 0.29 | 1.43 | 1.39 | 1.20 |
| **0.8** | **Day 21** | 0.12 | 0.13 | 0.15 | 1.35 | 1.15 | 1.17 |
| | **Day 63** | 1.00 | 1.00 | 1.00 | 0.53 | 0.78 | 0.67 |
| | **Fortnightly** | 0.90 | 0.61 | 0.86 | 0.56 | 0.95 | 0.79 |
| | **Weekly** | 1.00 | 0.71 | 0.96 | 0.16 | 0.86 | 0.66 |

#### 7 Additional figures

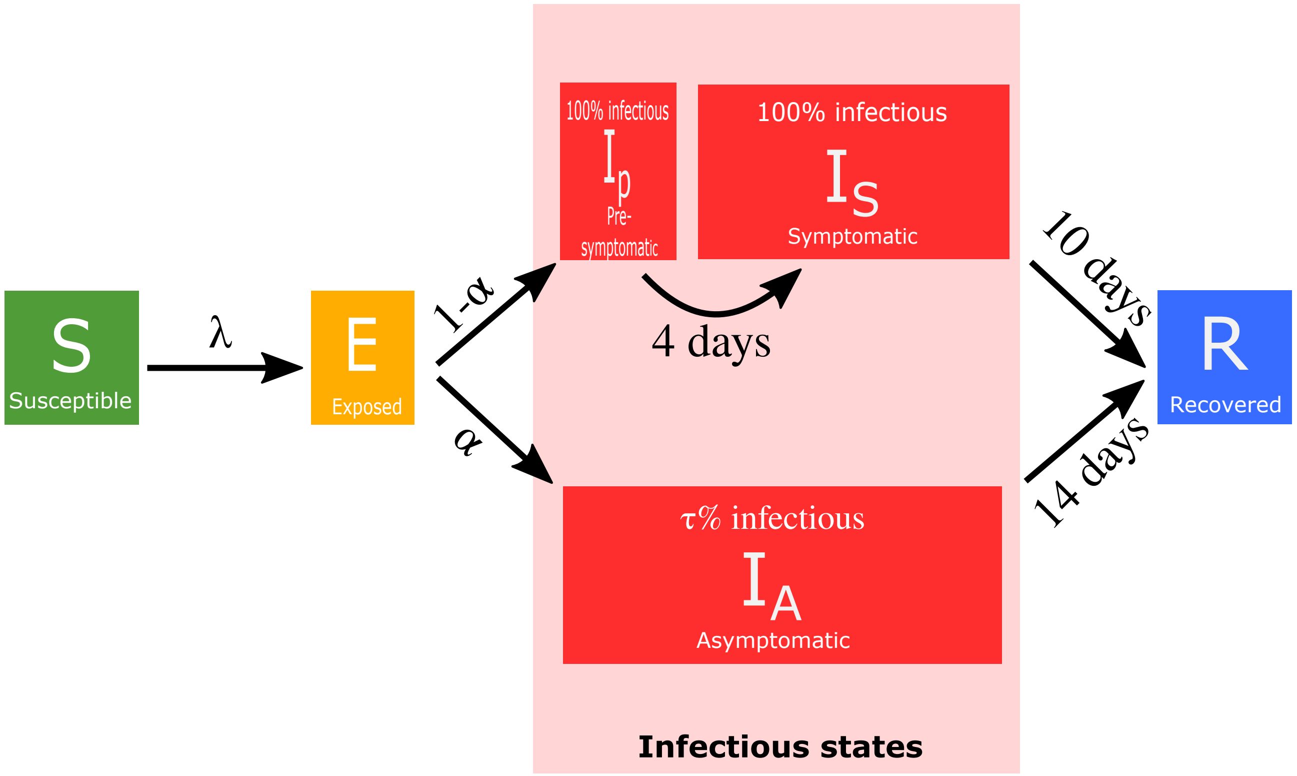


**Fig. S7: Epidemiological model disease states and transitions.** We stratified the population into susceptible ($S$), exposed ($E$, infected but not yet infectious), presymptomatic infectious ($I_p$), symptomatic infectious ($I_S$), asymptomatic infectious ($I_A$), and recovered ($R$) states. Solid lines denote transitions between disease states. A susceptible individual ($S$) becomes infected due to infectious pressure ($\lambda$) exerted by having contact with an infectious individuals, which can lead to onward transmission of the virus. Upon infection, individuals enter the exposed state ($E$). Upon leaving the exposed period, the individual enters the infectious state on one of two pathways: (i) with probability $\alpha \in [0.3, 0.7]$ they enter the asymptomatic state ($I_A$), with the parameter $\tau \in [0.5, 0.8]$ denoting the relative infectiousness of an asymptomatic versus a symptomatic case and presymptomatic cases that will become symptomatic; (ii) with probability 1-$\alpha$ they enter the presymptomatic state ($I_p$), before becoming symptomatic ($I_S$) after four days. Upon resolution of infection individuals move to the recovered ($R$) state. Infectious individuals leave the infectious state after 14 days, meaning infectious cases that went on to become symptomatic remain in the symptomatic state for 10 days.

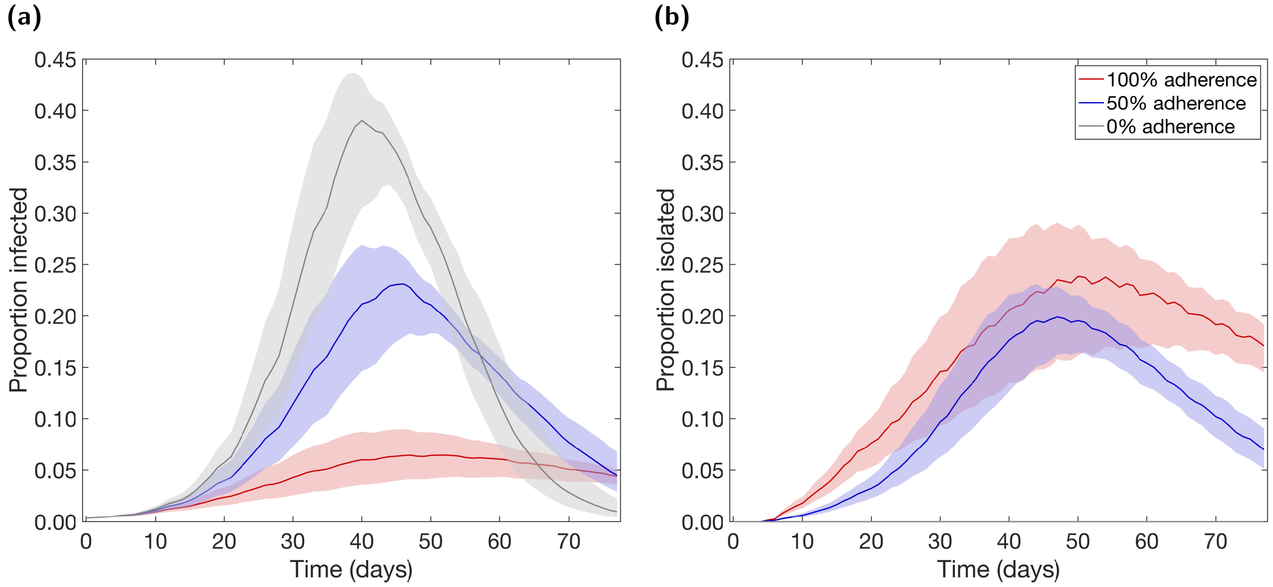


**Fig. S8: Temporal profiles of epidemiological measures over the autumn term under differing levels of adherence to nonpharmaceutical interventions.** Outputs produced from 1,000 simulations (with 20 runs per network, for 50 network realisations) for three levels of adherence to nonpharmaceutical interventions: 0% (grey), 50% (blue), 100% (red). Solid lines depict the median profile and shaded regions the 50% prediction interval. Patterns of infection prevalence and proportion of the student community in isolation demonstrated trade-offs between case numbers and the need for portions of the population to isolate. **(a)** Proportion infected. **(b)** Proportion isolated.

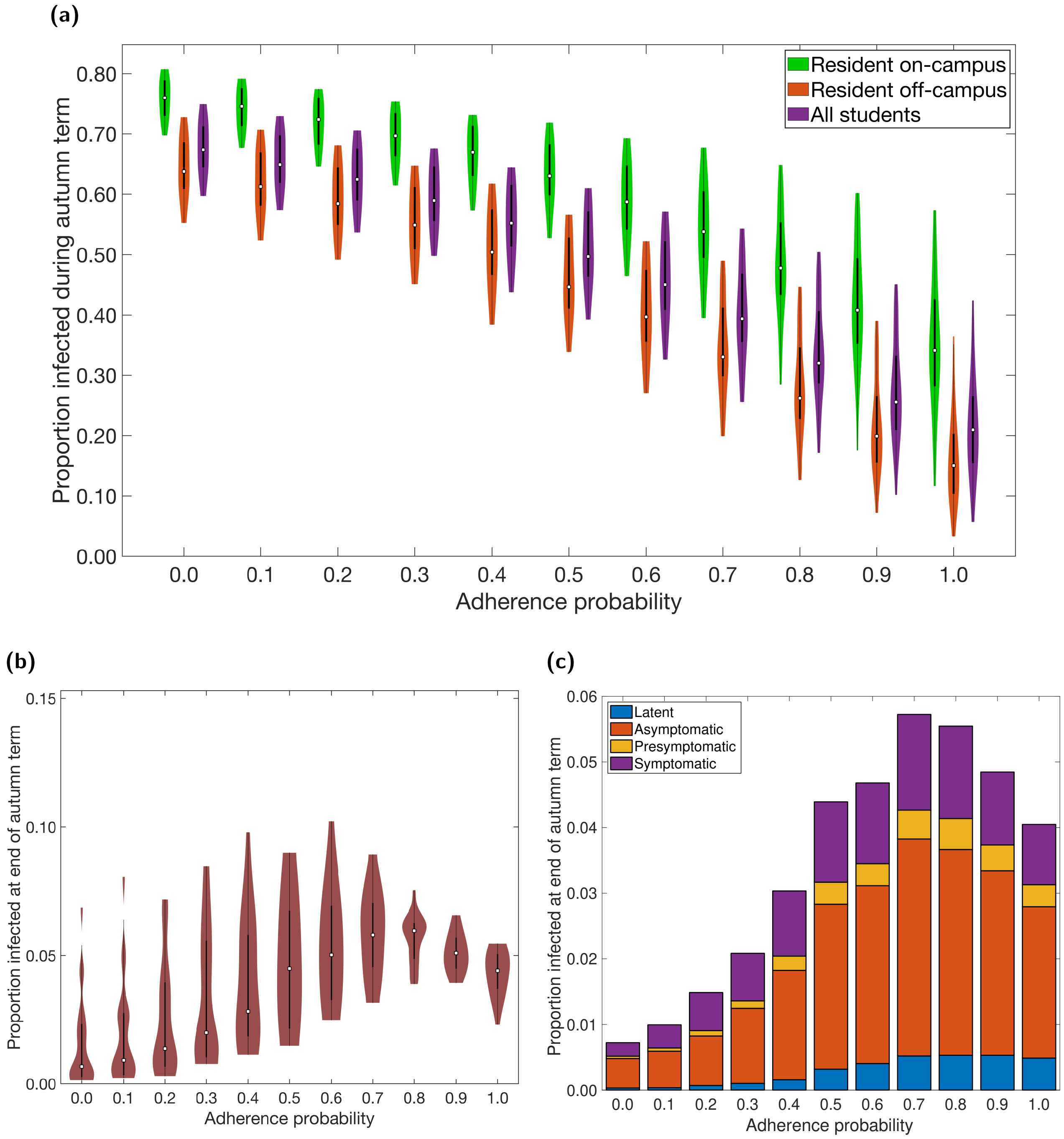


**Fig. S9: Infection epidemiological measures over the autumn term under differing levels of adherence to NPIs using a single network structure.** Outputs summarised from 20 simulations on a single network realisation. In all the violin plots, the white markers denote medians and solid black lines span the 25th to 75th percentiles. We observe a large amount of variability in outcomes, which results from differences in epidemiological properties between simulations runs, such as the distribution of initial infections, the asymptomatic probability, and the relative infectiousness of an asymptomatic case (all of which were randomly generated at the start of each simulation). **(a)** Over the duration of the autumn term, distributions relative to students resident on-campus only (green violin plots), students resident off-campus only (orange violin plots) and to the overall student population (purple violin plots) for proportion infected. **(b)** Proportion of students infected at the end of the autumn term. **(c)** Under each level of adherence, we display the median proportion of the student population in latent (blue), asymptomatic (orange), presymptomatic (yellow), symptomatic (purple) infected states at the end of the autumn term; for a given adherence value the height of the bar in panel (c) corresponds to the median point in panel (b).

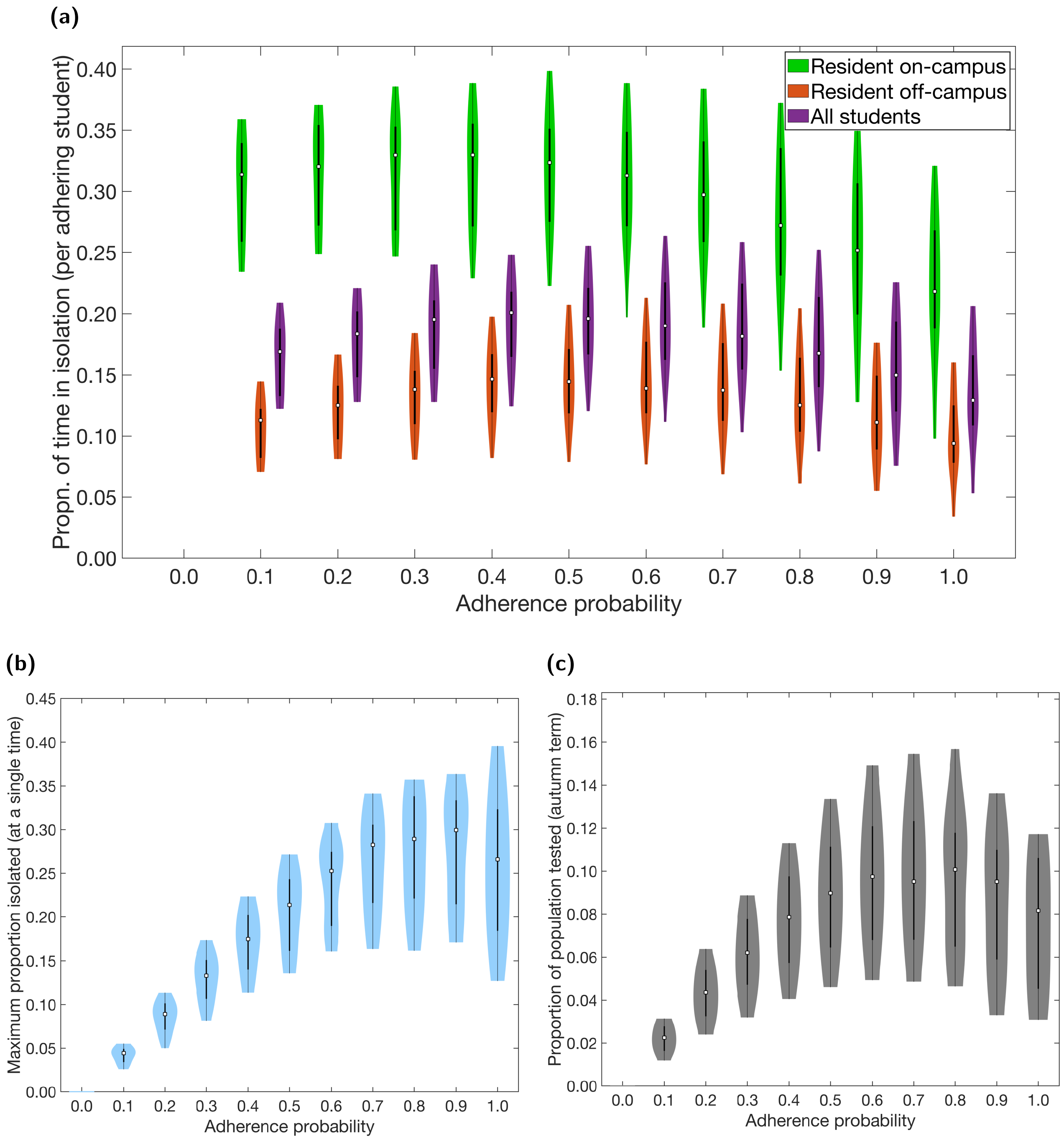


**Fig. S10: Non-infection epidemiological measures over the autumn term under differing levels of adherence to NPIs using a single network structure.** Outputs summarised from 20 simulations on a single network realisation. In all the violin plots, the white markers denote medians and solid black lines span the 25th to 75th percentiles. We observe a large amount of variability in outcomes, which results from differences in epidemiological properties between simulations runs, such as the distribution of initial infections, the asymptomatic probability, and the relative infectiousness of an asymptomatic case (all of which were randomly generated at the start of each simulation). **(a)** Over the duration of the autumn term, distributions relative to students resident on-campus only (green violin plots), students resident off-campus only (orange violin plots) and to the overall student population (purple violin plots) for proportion of time adhering students spend in isolation. **(b)** Maximum proportion of students isolated on a single day. **(c)** Proportion of population infected by SARS-CoV-2 and tested during the autumn term.

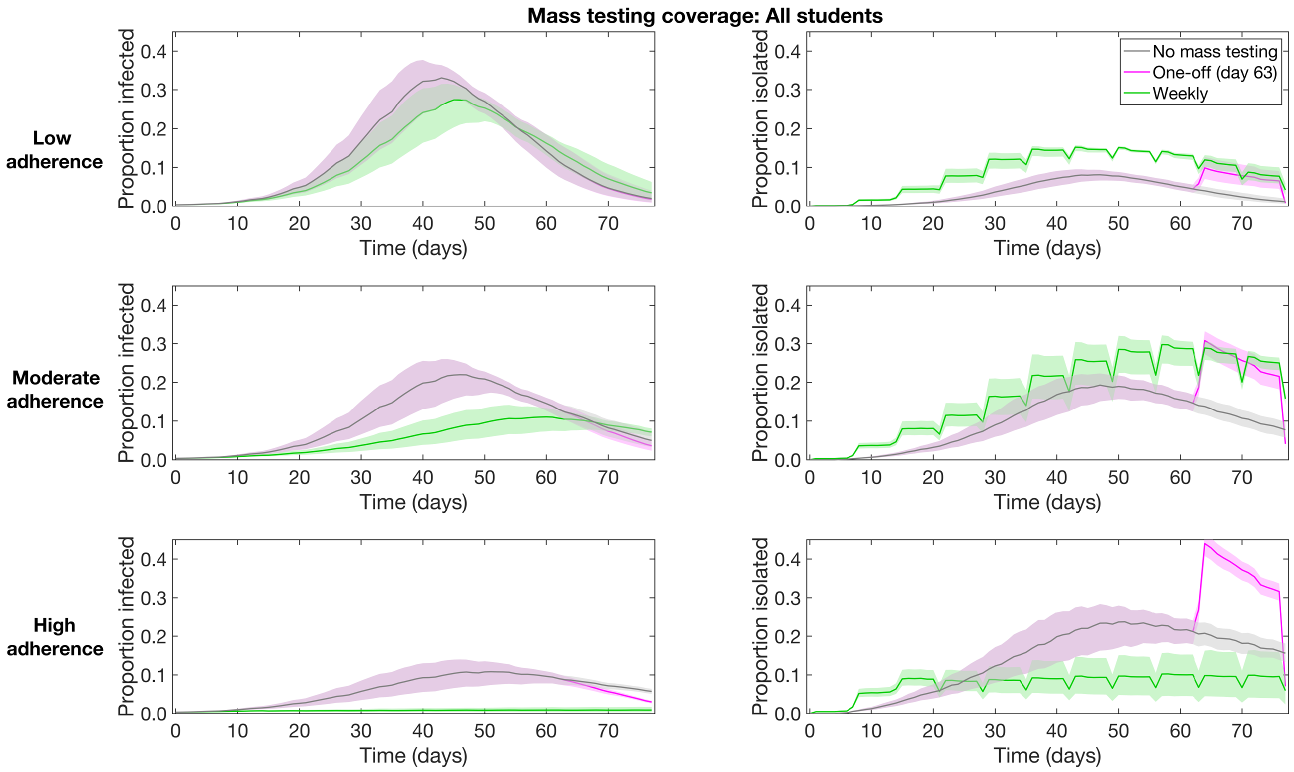


**Fig. S11: Infection prevalence and isolation temporal profiles of with mass testing of all students.** Outputs produced from 1,000 simulations (with 20 runs per network, for 50 network realisations) for no mass testing (grey), a single mass test instance on day 63 (magenta) and weekly mass testing (green). In all panels, solid lines depict the median profile and shaded regions the 50% prediction interval. We display outcomes of: **(Column one)** proportion infected; **(Column two)** proportion isolated. We used the following underlying probabilities of adhering to isolation measures: **(Row one)** 0.2; **(Row two)** 0.5; **(Row three)** 0.8. We found end of term prevalence was similar or lower for the one-off use of mass testing (on day 63) compared to weekly mass testing.

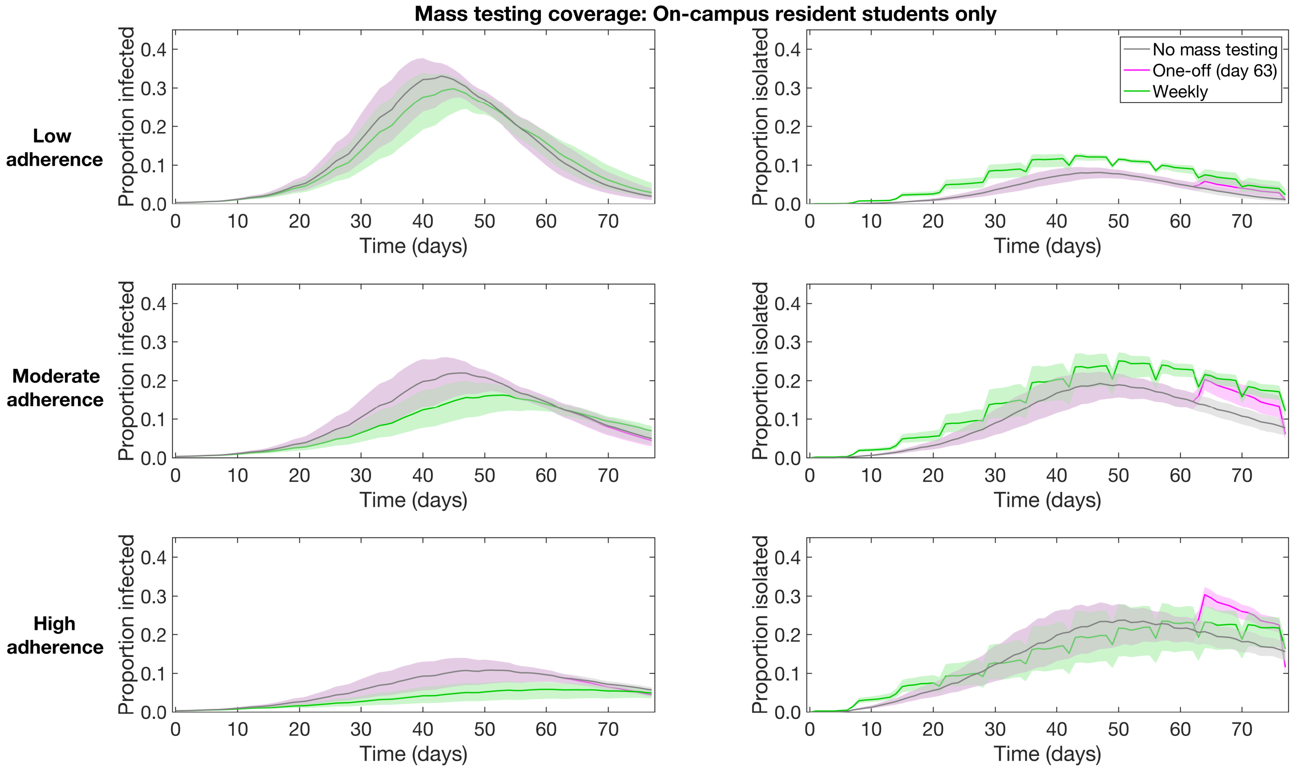


**Fig. S12: Infection prevalence and isolation temporal profiles of with mass testing of students resident on-campus.** Outputs produced from 1,000 simulations (with 20 runs per network, for 50 network realisations) for no mass testing (grey), a single mass test instance on day 63 (magenta) and weekly mass testing (green). In all panels, solid lines depict the median profile and shaded regions the 50% prediction interval. We display outcomes of: **(Column one)** proportion infected; **(Column two)** proportion isolated. We used the following underlying probabilities of adhering to isolation measures: **(Row one)** 0.2; **(Row two)** 0.5; **(Row three)** 0.8. We found end of term prevalence was similar or lower for the one-off use of mass testing (on day 63) compared to weekly mass testing.

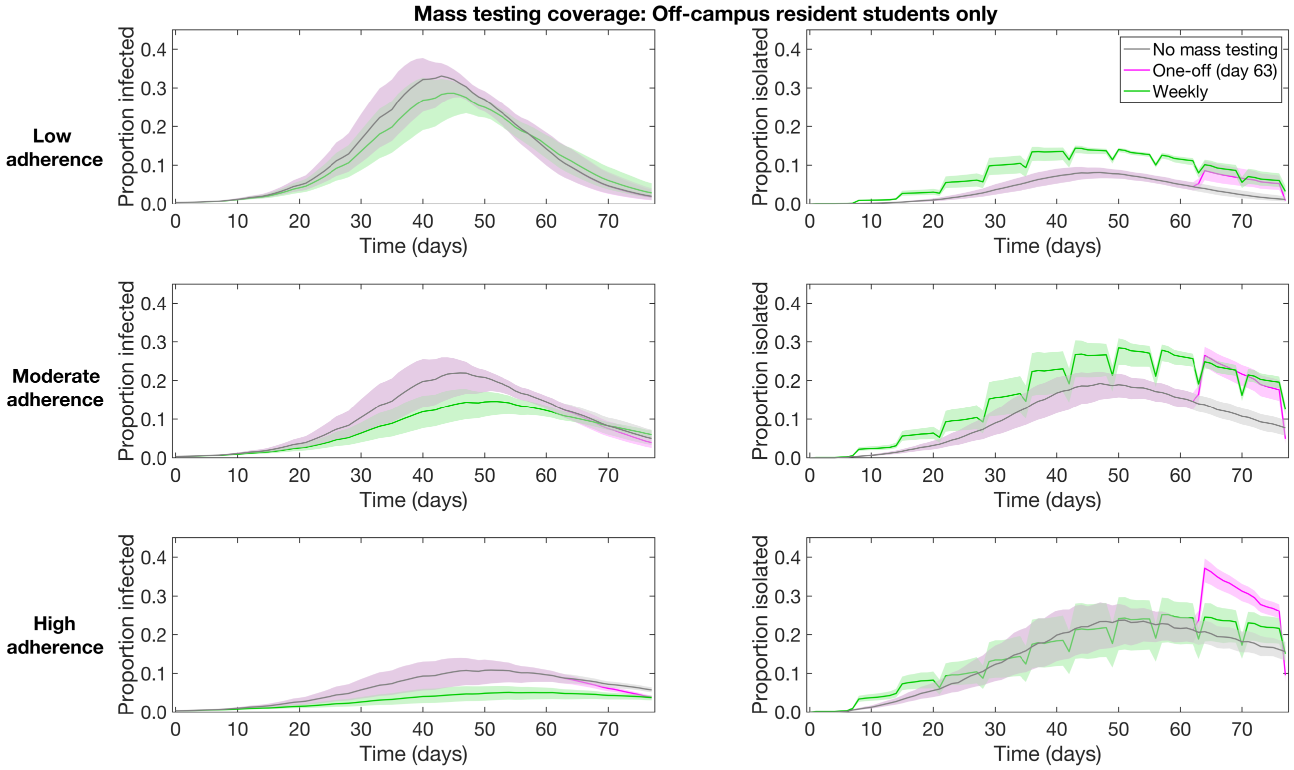


**Fig. S13: Infection prevalence and isolation temporal profiles of with mass testing of students resident off-campus.** Outputs produced from 1,000 simulations (with 20 runs per network, for 50 network realisations) for no mass testing (grey), a single mass test instance on day 63 (magenta) and weekly mass testing (green). In all panels, solid lines depict the median profile and shaded regions the 50% prediction interval. We display outcomes of: **(Column one)** proportion infected; **(Column two)** proportion isolated. We used the following underlying probabilities of adhering to isolation measures: **(Row one)** 0.2; **(Row two)** 0.5; **(Row three)** 0.8. We found end of term prevalence was similar or lower for the one-off use of mass testing (on day 63) compared to weekly mass testing.

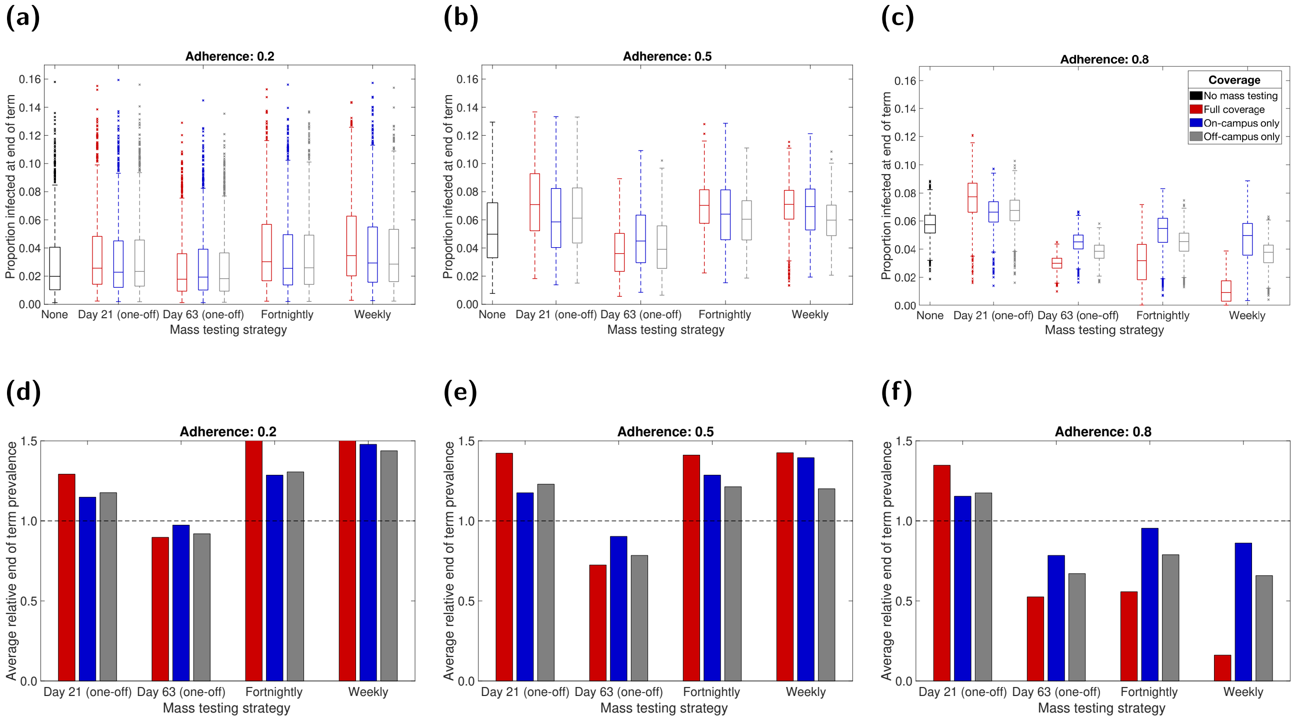


**Fig. S14: Proportion infected at the end of the academic term under differing mass testing strategies.** Mass testing was either not used (baseline scenario), a single instance took place at the end of week two or week eight of the academic term (corresponding to simulation day numbers 21 and 63, respectively), or regular mass testing was performed on a fortnightly or weekly basis. In each panel, we summarise outputs from 1000 simulations for mass testing covering all eligible students (red), on-campus only (blue), off-campus only (grey). Panels **(a-c)** compare the distributions of end of term infection prevalence with no mass testing (black) with outcomes for the various mass testing strategies. Panels **(d-f)** show the average scale of end of term infection prevalence for the given mass testing strategy relative to the scenario where no one-off mass testing event took place. The dashed line signifies parity between the scenarios. We used the following underlying probabilities of adhering to isolation measures: **(a,d)** 0.2; **(b,e)** 0.5; **(c,f)** 0.8. For a listing of values, see Tables S6-S7. End of term prevalence was minimised with a single one-off mass testing instance a fortnight before the term ended.
